# Multi-trait genome-wide association study identifies novel endometrial cancer risk loci that are associated with obesity or female testosterone levels

**DOI:** 10.1101/2021.08.01.21261455

**Authors:** Xuemin Wang, Pik Fang Kho, Dhanya Ramachandran, Cemsel Bafligil, Frederic Amant, Ellen L. Goode, Rodney J. Scott, Ian Tomlinson, D. Gareth Evans, Endometrial Cancer Association Consortium, Emma J. Crosbie, Thilo Dörk, Amanda B. Spurdle, Dylan M. Glubb, Tracy A. O’Mara

## Abstract

We have performed genetic correlation and Mendelian randomization analyses using publicly available genome-wide association study (GWAS) data to identify endometrial cancer risk factors. These and previously established risk factors of endometrial cancer were then included in a multi-trait Bayesian GWAS analysis to detect endometrial cancer susceptibility variants, identifying three novel loci (7q22.1, 8q24.3 and 16q12.2); two of which were replicated in an independent endometrial cancer GWAS dataset. These loci are hypothesized to affect endometrial cancer risk through altered sex-hormone levels or through effects on obesity. Consistent with this hypothesis, several genes with established roles in these pathways (*CYP11B1, CYP3A7, IRX3 and IRX5*) were prioritized as candidate endometrial cancer risk genes by interrogation of quantitative trait loci data and chromatin capture assays in endometrial cell lines. The findings of this study identify additional opportunities for hormone treatment and further support weight loss to reduce the risk of developing endometrial cancer.

**Statement of Significance:** This study prioritizes four genes related to testosterone and obesity as candidate endometrial cancer risk genes, as well as identifies additional opportunities for hormone treatment and further supports weight loss to reduce endometrial cancer risk.

## Introduction

Endometrial cancer is the fifth most common female cancer worldwide and the most common gynecological tumor in industrialized countries, accounting for over 380,000 new cases and nearly 90,000 deaths in 2018^1^. In addition, its prevalence and mortality rate are increasing in both high- and low-income countries^2-4^.

Identification of genetic susceptibility loci lays a foundation for the understanding of cancer etiology. Using case-control genome-wide association study (GWAS) data from multiple studies in the Endometrial Cancer Association Consortium (ECAC), we have identified 16 loci significantly associated with endometrial cancer risk^5-7^. Together, these risk loci are estimated to explain approximately a quarter of the familial relative risk attributable to common, readily-imputable variants^5^, indicating a large proportion of common endometrial cancer risk variants are still unidentified.

Endometrial cancer risk is influenced by many factors^5,8^. A series of Mendelian randomization studies, which use trait-associated genetic variants to infer causal relationships, have confirmed known and identified new endometrial cancer risk factors, including increased body mass index (BMI)^5,9,10^, early onset menarche^5,11^, low sex hormone binding globulin (SHBG), high testosterone^12^, increased serum estradiol^13^, increased fasting insulin^9^, and decreased low- (LDL) and increased high-density lipoproteins (HDL)^14^.

A recently developed multi-trait GWAS approach, the Bayesian GWAS (bGWAS) method^15^, uses priors derived from Mendelian randomization analyses of risk factors in a Bayesian framework to identify genetic variants associated with a trait of interest (focal trait). This approach not only offers an opportunity to identify novel genetic loci associated with a trait, but also provides an insight into which trait-associated loci for a given risk factor(s) could act through to affect the focal trait. The utility of this approach has been demonstrated in identifying new loci associated with lifespan^15-17^ and non-alcoholic fatty liver disease^18^.

In this study, we used GWAS summary-level data of diverse traits from the UK Biobank and other consortia to identify risk factors for endometrial cancer via genetic correlation and Mendelian randomization analyses. We implemented bGWAS to identify new endometrial cancer risk loci and to explore which risk factor pathways could mediate the effects of individual loci. Novel loci were assessed for replication in independent endometrial cancer GWAS datasets and functional interpretation provided to further our understanding of endometrial cancer etiology.

## Materials and Methods

### GWAS Summary Statistics

GWAS summary statistics of endometrial cancer risk were from the latest ECAC GWAS analysis (12,906 cases and 108,979 controls)^5^. To avoid bias due to overlapping sample sets, UK Biobank samples were removed from the ECAC summary statistics for Mendelian randomization and bGWAS analyses, resulting in 12,270 endometrial cancer cases and 46,126 controls. Secondary analyses were also performed using GWAS summary statistics restricted to endometrioid endometrial cancer histology only (8,758 cases and 46,126 controls).

GWAS summary statistics for potential risk factors were from publicly available resources, including the UK Biobank GWAS datasets^19^ analyzed by the Neale laboratory (http://www.nealelab.is/uk-biobank/), MAGIC (https://magicinvestigators.org/), GLGC (http://lipidgenetics.org/), the GWAS Catalog (https://www.ebi.ac.uk/gwas/), BCAC (http://bcac.ccge.medschl.cam.ac.uk/), and the ReproGen Consortium (https://www.reprogen.org). Given endometrial cancer occurs only in women, where available we used female-stratified GWAS data. Sex-combined GWAS summary statistics were used for traits where female-stratified GWAS summary statistics were not available. Duplicated traits were excluded manually. Further information for trait GWAS summary statistics used in this study is provided in **Supplementary Table S1**.

### Genetic Correlation

Genetic correlations between endometrial cancer risk and available traits were estimated using publicly available GWAS summary statistics and LD score regression (LDSC version 1.0.1)^20^. Significant correlations were defined as those with a false discovery rate (FDR) < 0.01. Since BMI is established to be the strongest risk factor for endometrial cancer^10^ and many traits are correlated with BMI, we additionally estimated the genetic correlation between potential risk factors and endometrial cancer risk after adjusting for BMI. Adjustment for BMI was conducted by fitting the female-stratified UK Biobank GWAS summary statistics of BMI as a covariate of the analysis of endometrial cancer using GWAS summary-level data using the mtCOJO method^21^ implemented in GCTA (version 1.91.7beta)^22^. Linkage disequilibrium (LD) was estimated using individual genotypic data from a random set of 10,000 unrelated individuals from the UK Biobank cohort (https://www.ukbiobank.ac.uk/) selected by Kho et al. (2020)^14^.

### Mendelian Randomization Analysis

Traits that remained significantly genetically correlated with endometrial cancer after adjusting for BMI were assessed for endometrial cancer causality using two-sample Mendelian randomization analyses. Independent genetic variants robustly associated with individual traits (P < 5 × 10^−8^) were selected as instrumental variables using the stepwise model selection procedure implemented by the GCTA-COJO function^23^. Default settings were used apart from --cojo-wind, which was set to 1000. A/T or C/G allelic variants with minor allele frequency larger than 0.42 were excluded from analyses due to the ambiguity into the identity of the effect allele in the exposure and outcome^24^. Mendelian randomization analyses were performed using the TwoSampleMR R package (version 0.5.5)^24^. As the GWAS analyses of endometrial, breast, and ovarian cancers included shared control participants, the latter two cancers were excluded from the Mendelian randomization analyses.

### Bayesian Genome-wide Association Study (bGWAS)

Multi-trait GWAS analysis of endometrial cancer was conducted using the bGWAS framework implemented in R^15^. Traits included in the bGWAS analysis can be found in **Supplementary Table 1**. These included traits found to be causally associated with endometrial cancer risk in this study plus risk factors reported by previous Mendelian randomization analyses (i.e. BMI^5,9,10^, testosterone^12^, estradiol^13^, age at menarche^5,11^, HDL-cholesterol^14^, and LDL-cholesterol^14^). Breast and ovarian cancers were excluded from this analysis due overlapping control participants with the endometrial cancer GWAS dataset as mentioned above.

**Table 1.** Traits significantly genetically correlated with endometrial cancer risk after adjustment for BMI and Mendelian randomisation analysis. **Abbreviations:** BMI - body mass index; r_G_ - genetic correlations intercept; SE - standard error; SHBG - sex hormone binding globulin nsnps - number of variants included in Mendelian randomisation analysis; OR - odds ratio; CI - confidence interval

| Phenotype | Genetic correlation results before adjusting for BMI |  |  | Genetic correlation results after adjusting for BMI |  |  | Inverse variance weighted Mendelian randomisation analysis |  |  |
| --- | --- | --- | --- | --- | --- | --- | --- | --- | --- |
| | $r_G$ | SE | P-value | $r_G$ | SE | P-value | nsnps | OR (95% CI) | P-value |
| Increased arm fat mass (left) | 0.58 | 0.07 | 6.28E-17 | 0.17 | 0.05 | 5.0E-04 | 137 | 1.67 (1.44 -1.95) | 2.8E-11 |
| Increased arm fat mass (right) | 0.58 | 0.07 | 7.54E-17 | 0.17 | 0.05 | 5.0E-04 | 143 | 1.56 (1.35 -1.81) | 2.8E-09 |
| Increased arm predicted mass (left) | 0.46 | 0.06 | 2.91E-14 | 0.18 | 0.05 | 4.0E-04 | 253 | 1.26 (1.12 -1.41) | 8.9E-05 |
| Increased basal metabolic rate | 0.47 | 0.06 | 5.50E-15 | 0.18 | 0.05 | 2.0E-04 | 274 | 1.29 (1.17 -1.44) | 1.1E-06 |
| Increased body fat percentage | 0.54 | 0.07 | 2.81E-15 | 0.17 | 0.05 | 3.0E-04 | 130 | 1.52 (1.30 -1.79) | 2.5E-07 |
| Breast Cancer | 0.25 | 0.05 | 3.20E-06 | 0.29 | 0.06 | 3.6E-07 | not assessed due to sample overlap |  |  |
| Increased hip circumference | 0.56 | 0.07 | 5.46E-16 | 0.19 | 0.05 | 1.0E-04 | 150 | 1.46 (1.23 -1.74) | 2.02E-05 |
| Increased leg fat mass (left) | 0.58 | 0.07 | 6.78E-17 | 0.17 | 0.05 | 4.0E-04 | 144 | 1.59 (1.38 -1.83) | 7.19E-11 |
| Increased leg fat mass (right) | 0.58 | 0.07 | 6.79E-17 | 0.17 | 0.05 | 4.0E-04 | 143 | 1.59 (1.37 -1.85) | 1.00E-09 |
| Increased leg fat ratio | -0.29 | 0.05 | 3.13E-09 | -0.19 | 0.05 | 2.0E-04 | 55 | 0.95 (0.78 -1.16) | 0.64 |
| Increased leg fat-free mass (left) | 0.46 | 0.06 | 2.03E-14 | 0.19 | 0.05 | 2.0E-04 | 280 | 1.23 (1.11 -1.37) | 5.51E-05 |
| Increased leg fat-free mass (right) | 0.44 | 0.06 | 6.52E-14 | 0.18 | 0.05 | 3.0E-04 | 292 | 1.24 (1.12 -1.36) | 2.17E-05 |
| Increased leg predicted mass (left) | 0.46 | 0.06 | 2.68E-14 | 0.19 | 0.05 | 2.0E-04 | 278 | 1.25 (1.13 -1.38) | 2.48E-05 |
| Increased leg predicted mass (right) | 0.44 | 0.06 | 9.25E-14 | 0.18 | 0.05 | 4.0E-04 | 288 | 1.22 (1.11 -1.35) | 3.29E-05 |
| Increased metabolic syndrome risk | 0.46 | 0.06 | 1.06E-15 | 0.23 | 0.06 | 1.0E-04 | 76 | 1.03 (0.95 -1.12) | 0.48 |
| Ovarian cancer | 0.42 | 0.10 | 3.55E-05 | 0.47 | 0.12 | 6.5E-05 | not assessed due to sample overlap |  |  |
| Late relative age of first facial hair | -0.29 | 0.06 | 1.39E-06 | -0.23 | 0.07 | 7.0E-04 | 86 | 0.70 (0.52 -0.95) | 0.02 |
| Increased SHBG levels | -0.46 | 0.07 | 5.90E-10 | -0.26 | 0.06 | 3.8E-05 | 165 | 0.85 (0.79 -0.92) | 2.41E-05 |
| Increased trunk fat mass | 0.55 | 0.07 | 1.82E-16 | 0.21 | 0.05 | 1.2E-05 | 156 | 1.45 (1.25 -1.69) | 1.47E-06 |
| Increased trunk fat percentage | 0.52 | 0.07 | 1.01E-14 | 0.20 | 0.05 | 5.4E-05 | 141 | 1.48 (1.26 -1.73) | 1.95E-06 |
| Increased weight | 0.56 | 0.07 | 2.07E-17 | 0.20 | 0.05 | 2.8E-05 | 192 | 1.39 (1.22 -1.59) | 8.22E-07 |
| Increased whole body fat mass | 0.57 | 0.07 | 4.64E-17 | 0.20 | 0.05 | 4.6E-05 | 149 | 1.52 (1.31 -1.76) | 5.87E-08 |
| Increased whole body fat-free mass | 0.41 | 0.06 | 5.05E-13 | 0.17 | 0.05 | 7.0E-04 | 300 | 1.21 (1.10 -1.33) | 1.36E-04 |
| Increased whole body water mass | 0.41 | 0.06 | 4.05E-13 | 0.17 | 0.05 | 7.0E-04 | 301 | 1.20 (1.09 -1.33) | 1.95E-04 |

The bGWAS method involves two main steps: (1) identification of independent risk factors that jointly affect endometrial cancer risk; and (2) determination of variant-trait associations by multi-trait GWAS using risk factors identified in step 1. Specifically, in the first step, univariate regression analyses were conducted to identify endometrial cancer risk factors using instrumental variables associated with traits (P < 1 × 10^−5^). Instrumental variables were considered independent if their physical distances were over 500 kb. Risk factors identified by univariate regression analyses were subsequently included in a multivariate Mendelian randomization analysis to determine independent risk factors of endometrial cancer via a stepwise selection procedure. In the second step, prior effects of individual variants were estimated as the sum of the products of the variant effects on individual risk factors and the causal effects of individual risk factors on endometrial cancer risk. Bayesian factors (BFs) and the corresponding p-value (P_BF_) were calculated. Significant associations identified based on BFs represent variants exerting their effects on endometrial cancer risk through risk factors included in the analysis. Direct and posterior effects and their corresponding p-values were also estimated. Variants showing a significant association by direct effects are likely to affect endometrial cancer risk directly or through other risk factors not included in the analysis; whereas variants displaying large prior effects, driven by one or more risk factors, could show significant posterior effects. A genome-wide significance threshold (P < 5 × 10^−8^) was used to identify significant variant-trait associations. We highlighted potential endometrial cancer risk loci if a genome-wide significant signal was observed more than 500 kb from a known endometrial cancer risk locus.

To identify potential subtype-specific loci, we performed an additional bGWAS analysis using the endometrioid endometrial cancer risk GWAS summary statistics. This was performed as above, apart from the threshold used for instrumental variable inclusion (P < 1 × 10^−6^, instead of 1 × 10^−5^) because of the improved correlation between prior and observed effects of genetic variants (**Supplementary Table S2**).

To minimize false positives, following bGWAS analysis, the following criteria were applied to restrict output variants: (1) concordant direction of effect on endometrial cancer risk in this study and in the most recent ECAC GWAS^5^, and (2) a nominally significant association (P < 0.05) with endometrial cancer risk in the most recent ECAC GWAS^5^.

### Endometrial Cancer GWAS Replication

To construct a replication set and validate our findings from the bGWAS analysis, we downloaded publicly available endometrial cancer GWAS summary statistics which were not included in the ECAC GWAS data^5^. These were from the Finnish biobank study, FinnGen (data freeze 2, 566 cases and 75,822 controls; http://r2.finngen.fi/), and from the Japanese Biobank study^25^ (999 cases and 89,731 controls; http://jenger.riken.jp/en/). Endometrial cancer GWAS summary statistics from the Manchester cohort (UK) were provided by collaboration (560 cases and 1,202 controls)^26,27^. Since the UK Biobank endometrial cancer case-control samples were excluded from the bGWAS analysis, we additionally included UK Biobank samples in our replication set. In total, 1,866 endometrial cancer cases were identified from UK Biobank (phenotype data accessed November 2020) using ICD10 code C54 in data fields 40006, 41270 and 41202. Genotypes for these women were extracted, with 18,660 non-related, randomly-selected women participants. Case-control GWAS analyses for this subset of UK Biobank participants were performed by REGENIE^28^, using a logistic mixed model including the genetic relationship matrix as a random effect to account for cryptic relatedness and population stratification. The top ten principal components and genotyping array were included as covariates in the model. Before analysis, quality control was performed to remove variants with a minor allele frequency < 1%, minor allele count < 100, genotype missingness > 10% and those that deviate from Hardy-Weinberg equilibrium (P_HWE_ < 1×10^−15^).

Summary statistics for each set were harmonized to the same genomic build (hg19) and variants with low minor allele frequency (< 1%) or low imputation quality (R^2^ < 0.3) were removed. The summary statistics from the four sets were combined in a fixed-effects, inverse-variance weighted meta-analysis by METAL^29^ (version release 2020-05-05), adjusting each set for genomic control.

### In silico Functional Analysis

For each locus identified using BFs, candidate causal endometrial cancer risk variants were defined as those with a P_BF_ 100:1 log likelihood ratio with the lead risk variant and located within ±500 kb of the lead variant. Candidate causal risk variants were interrogated with the Qtlizer tool^30^ to identify variants that were lead expression QTLs in GTEx tissues^31^. Genes encompassing candidate causal risk variants were browsed in the GTEx web portal^31^ to identify lead splicing QTLs among the candidate causal set of risk variants. To identify candidate target genes, candidate causal variants were intersected with promoter-associated H3K27Ac HiChIP chromatin loops captured from normal immortalized (E6E7hTERT) and tumoral (ARK1, Ishikawa, and JHUEM-14) endometrial cell lines^32^. To identify transcription factor binding sites, 10 bases flanking the variants of interest (effect allele or other allele) were taken to predict allele specific changes to transcription factor binding. Sequence based predictions were performed by TOMTOM^33^ (MEMESuitev5.3.3, mapped with the HOCOMOCOv11_full_HUMAN database). Spearman correlations between transcription factor gene expression and *CYP3A7* were performed in GTEx tissues using GEPIA2^34^.

## Results

### Genetic correlations with endometrial cancer risk

A flowchart overview of the study is presented in **Figure 1**. We assessed genetic correlations between 2,747 phenotypes and endometrial cancer risk (**Supplementary Table S1**). Endometrial cancer risk significantly correlated with 67 traits (false discovery rate; FDR < 0.01), with genetic correlations ranging from -0.46 to 0.59 (**Figure 2A**; **Supplementary Table S3**). Most traits clustered into seven groups (**Supplementary Figure S1A**): anthropometric traits such as fat or fat-free mass of limbs and body (13 traits), BMI (18 traits) and impedance measurements (5 traits); blood pressure-related traits (5 traits); pubertal traits (3 traits); ankle spacing (3 traits); and lipoproteins (2 traits).

**Figure 1.**
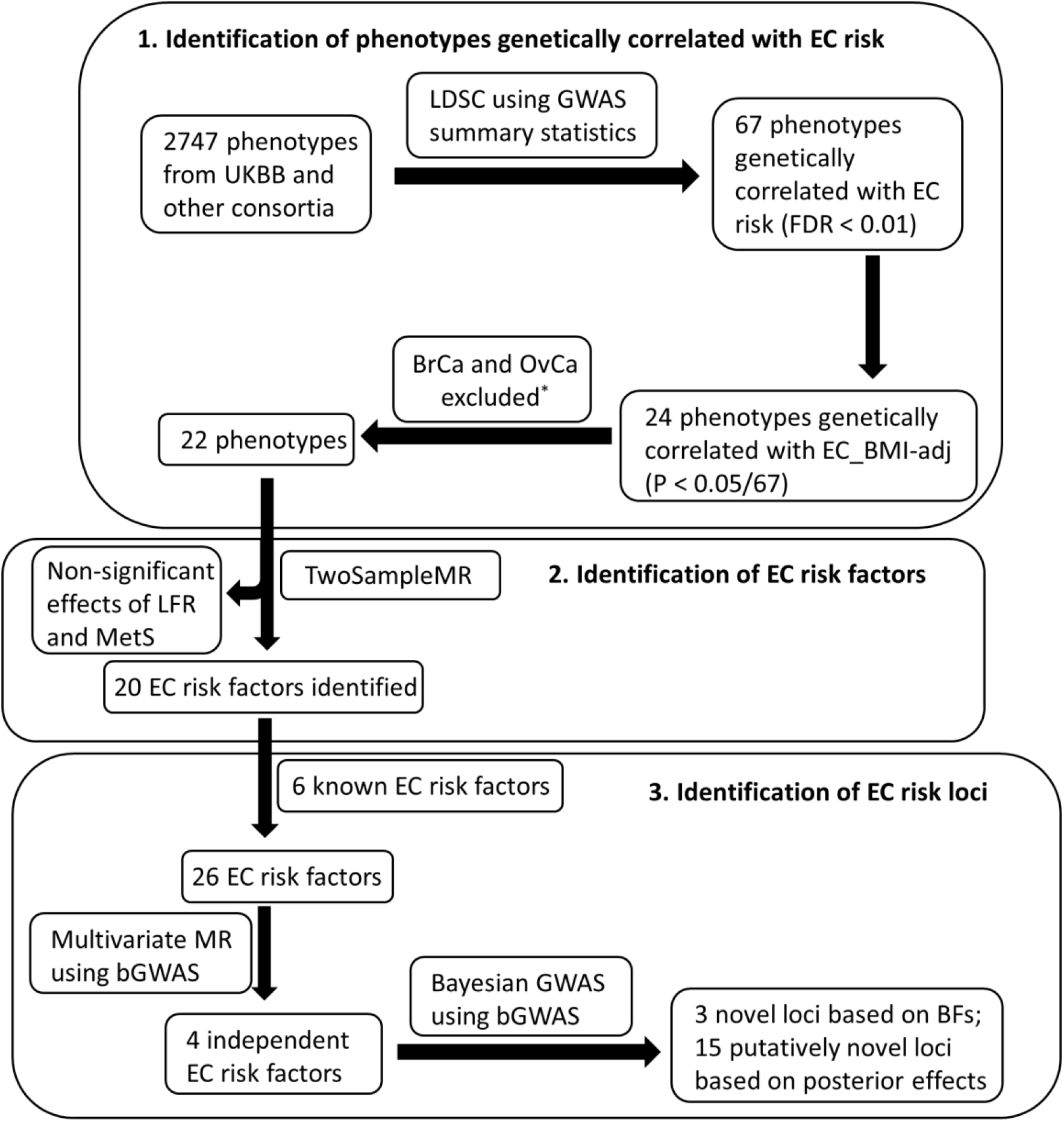
Schematic overview of the study. EC – endometrial cancer; UKBB – UK Biobank; EC_BMI-adj – endometrial cancer adjusted for BMI; FDR – false discovery rate; LDSR – LD score regression; MR – Mendelian randomization analysis; LFR – leg fat ratio; MetS – metabolic syndrome; bGWAS – Bayesian GWAS analysis; BFs – Bayesian factors. *breast cancer (BrCa) and ovarian cancer (OvCa) were excluded from univariate Mendelian randomization analyses due to overlapping control participants with the endometrial cancer GWAS.

**Figure 2.**
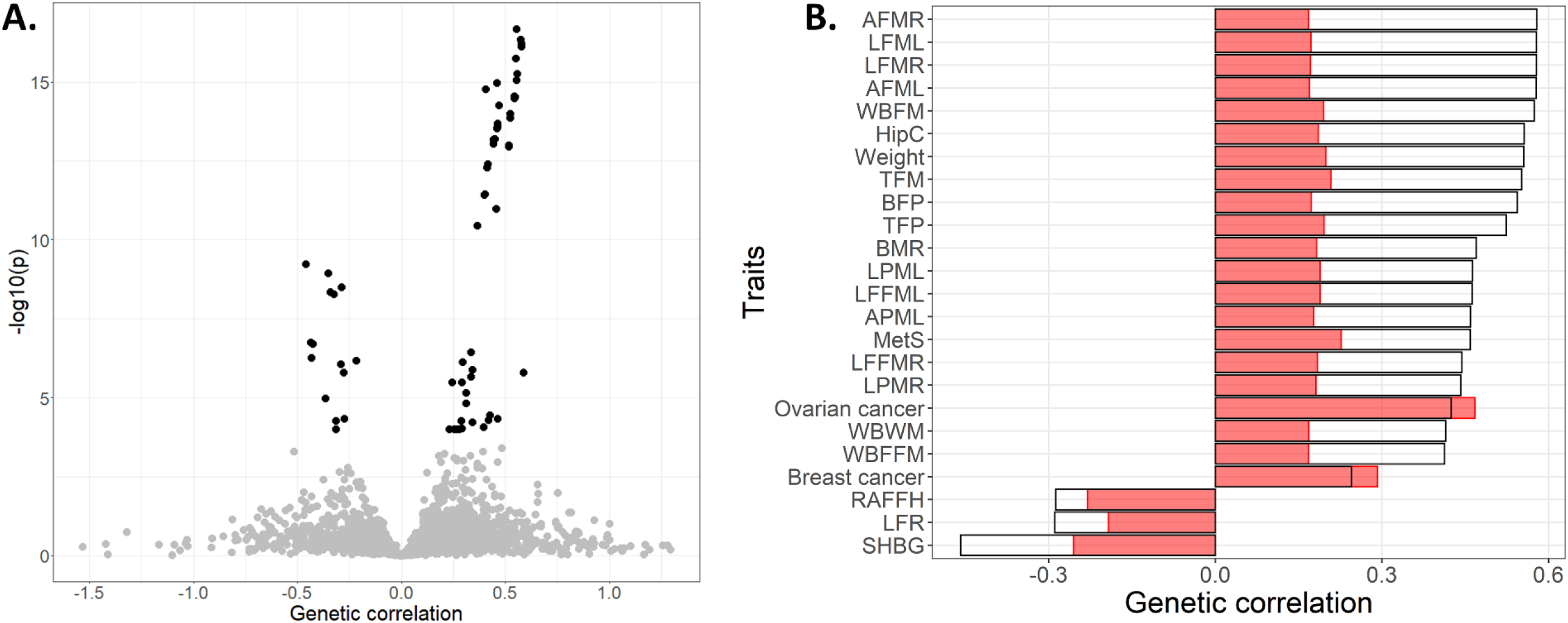
Genetic correlations between traits and endometrial cancer risk. **A**. Volcano plot of genetic correlations between traits and endometrial cancer risk. Black points represent 67 traits, which had a genetic correlation to endometrial cancer at false discovery rate (FDR) < 0.01. **B**. Bar plot of genetic correlation of 24 traits significantly correlated with endometrial cancer with or without adjustment for body mass index (BMI). Height of empty bars indicates the magnitude of genetic correlation to endometrial cancer without adjustment for BMI, whereas height of red bars indicates the magnitude of correlations to endometrial cancer adjusted for BMI. Abbreviations can be found in **Supplementary Table S3**.

After adjusting for BMI, endometrial cancer risk significantly correlated with 24 traits (P < 7.46 × 10^−4^; Bonferroni threshold = 0.05/67 significantly correlated traits before BMI adjustment) (**Table 1**; **Figure 2B**).The magnitude and significance level of these adjusted correlations were attenuated, with the exception of breast and ovarian cancers, which were slightly strengthened. The majority of the 24 traits (75%) related to anthropometric measurements and clearly clustered to the same group (**Supplementary Figure S1B**).

### Identification of endometrial cancer risk factors

Although breast and ovarian cancers were genetically correlated with endometrial cancer risk after adjustment for BMI, they were excluded from Two-sample Mendelian randomization analyses due to overlapping control participants with the endometrial cancer GWAS violating the principles of this analysis. Of the remaining 22 BMI-adjusted correlated traits, all but leg fat ratio and metabolic syndrome displayed significant effects on endometrial cancer risk by inverse variance weight (IVW) Mendelian randomization analyses (**Table 1**; **Supplementary Table S4**). Two of the 20 traits were found to reduce endometrial cancer risk: later relative age of first facial hair (OR 0.70; 95% CI 0.52-0.95; P = 0.02), and increased sex hormone binding globulin (SHBG) levels (OR 0.85; 95% CI 0.79-0.92; P = 2.41 × 10^−5^). The remaining 18 traits were found to increase endometrial cancer risk, with odd ratios varying from 1.20 (95% CI 1.09-1.33; p = 1.95 × 10^−4^) for increased whole body water mass to 1.67 (95% CI 1.44-1.95; p = 2.83 × 10^−11^) for increased left arm fat mass. Horizontal pleiotropy tests showed that Mendelian randomization results were not confounded by horizontal pleiotropy (MR-Egger intercept P-value > 0.05) (**Supplementary Table S4**).

After BMI adjustment, increased SHBG level was the only trait that maintained a significant effect reducing endometrial cancer risk (OR 0.86; 95% CI 0.80-0.93; P = 1.42 × 10^−4^) (**Supplementary Table S4**). The effect was similar to that before BMI adjustment, indicating the protective effect of increased SHBG levels on reducing endometrial cancer risk is independent of BMI.

### Identification of independent endometrial cancer risk factors and multi-trait GWAS

Together with the 20 risk factors identified above, six additional endometrial cancer risk factors reported previously based on genetic analysis (i.e. BMI, testosterone levels, estradiol levels, age at menarche, HDL-cholesterol levels, and LDL-cholesterol levels) were included in the multi-trait GWAS analysis of endometrial cancer (**Supplementary Table S1**). Multivariate Mendelian randomization analyses found four independent risk factors jointly displaying a significant effect on endometrial cancer risk: testosterone levels, SHBG levels, BMI, and age at menarche (**Supplementary Figure S2A**). We additionally performed multivariate Mendelian randomization analyses restricting to endometrioid endometrial cancer only and found testosterone, SHBG levels, BMI, and relative age of first facial hair to independently affected endometrioid endometrial cancer risk (**Supplementary Figure S2B**). This was not substantially different from the primary analysis of endometrial cancer risk as relative age of first facial hair (found to affect endometrioid endometrial cancer risk) belonged to the same cluster of traits as age at menarche (found to affect endometrial cancer risk) (**Supplementary Figure S3**).

Multi-trait GWAS analyses were conducted by bGWAS using priors constructed from the above endometrial cancer risk factors. The primary bGWAS analysis is based on Bayes factors (BFs), and identifies variants significantly associated with endometrial cancer predisposition through the risk factors included in the analysis. We also assessed direct effects from bGWAS analysis, which identifies variants likely to affect endometrial cancer risk directly, or through risk factors not included in the multi-trait GWAS analysis. As a secondary analysis, we considered variants identified by bGWAS posterior effects, which identifies associations displaying very large prior effects. These are associations which are largely driven by the relationship between the variant and at least one risk factor.

Our primary multi-trait BF GWAS analysis detected five loci associated with endometrial cancer risk (P_BF_ < 5 × 10^−8^) (**Figure 3A**; **Table 2**), two of which were known risk loci (12q24.12 and 15q15.1). The other three loci (7q22.1, 8q24.3, and 16q12.2) have not previously been reported as genome-wide significant endometrial cancer risk loci, although 7q22.1 has been reported as a sub-genome-wide significant endometrial cancer risk locus^5^. The four risk factors all contributed prior effects on endometrial cancer risk at the five loci (**Figure 3B, Table 2**), with three risk loci (7q22.1, 8q24.3, 15q15.1) significantly associated with testosterone levels. The novel endometrial cancer risk locus, 16q12.2, was associated with BMI, SHBG levels, and age at menarche (**Figure 3B**; **Table 2**). Similar results were found in multi-trait analyses for endometrioid endometrial cancer, with the same three novel loci detected (**Supplementary Figure S4**; **Table 2**).

**Table 2.** Genome-wide significant variant associations with risk of endometrial cancer based on Bayes Factors by bGWAS. Abbreviations: EA - effect allele; OA - other allele; Z-score - association estimate; µ - prior effect estimate; SE - standard error of µ; BF - Bayes Factor; P_BF_ - Bayes Factor p-value

| Region | rsid | nearby or candidate gene<sup>a</sup> | chr:pos (hg19) | EA | OA | All histologies |  |  |  |  | Endometrioid histology |  |  |  |  | Z-scores for risk factors included in multivariate GWAS |  |  |  |
| --- | --- | --- | --- | --- | --- | --- | --- | --- | --- | --- | --- | --- | --- | --- | --- | --- | --- | --- | --- |
|  |  |  |  |  |  | Z-score | μ | SE | BF | P<sub>BF</sub> | Z-score | μ | SE | BF | P<sub>BF</sub> | BMI | Testosterone levels | SHBG levels | Age of menarche |
| Novel endometrial cancer risk loci |  |  |  |  |  |  |  |  |  |  |  |  |  |  |  |  |  |  |  |
| 7q22.1 | rs34670419 | ZKSCAN5, CYP3A7 | 7:99130834 | T | G | -4.61 | -1.89 | 0.43 | 1671.23 | 1.23E-12 | -3.94 | -1.52 | 0.49 | 200.29 | 3.40E-10 | -3.45 | -30.71 | -4.24 | 2.41 |
| 8q24.3 | rs10103405 | CYP11B1 | 8:143968631 | C | T | -4.48 | -0.81 | 0.17 | 32.55 | 4.55E-08 | -4.74 | -0.75 | 0.19 | 33.72 | 4.50E-08 | -0.20 | -10.57 | -0.82 | 0.17 |
| 16q12.2 | rs56094641 | FTO, IRX3, IRX5 | 16:53806453 | G | A | 3.53 | 1.45 | 0.23 | 63.82 | 7.69E-09 | 3.48 | 1.31 | 0.28 | 46.65 | 1.84E-08 | 20.98 | 0.33 | -6.56 | -10.91 |
| Previously known endometrial cancer risk loci |  |  |  |  |  |  |  |  |  |  |  |  |  |  |  |  |  |  |  |
| 12q24.12 | rs7310615 | SH2B3 | 12:111865049 | G | C | 6.25 | 0.57 | 0.12 | 38.26 | 2.96E-08 | 5.73 | 0.50 | 0.13 | 19.71 | 2.03E-07 | 3.18 | 3.24 | -1.32 | -4.11 |
| 15q15.1 | rs998713 | BMF | 15:40378467 | G | A | 5.71 | 0.87 | 0.18 | 145.92 | 8.94E-10 | 4.45 | 0.70 | 0.20 | 22.77 | 1.35E-07 | -0.48 | 11.69 | 0.95 | -2.46 |

<sup>a</sup>Bolded genes are candidate genes identified from analyses described in this study or reported from O'Mara et al (2019)

**Figure 3.**
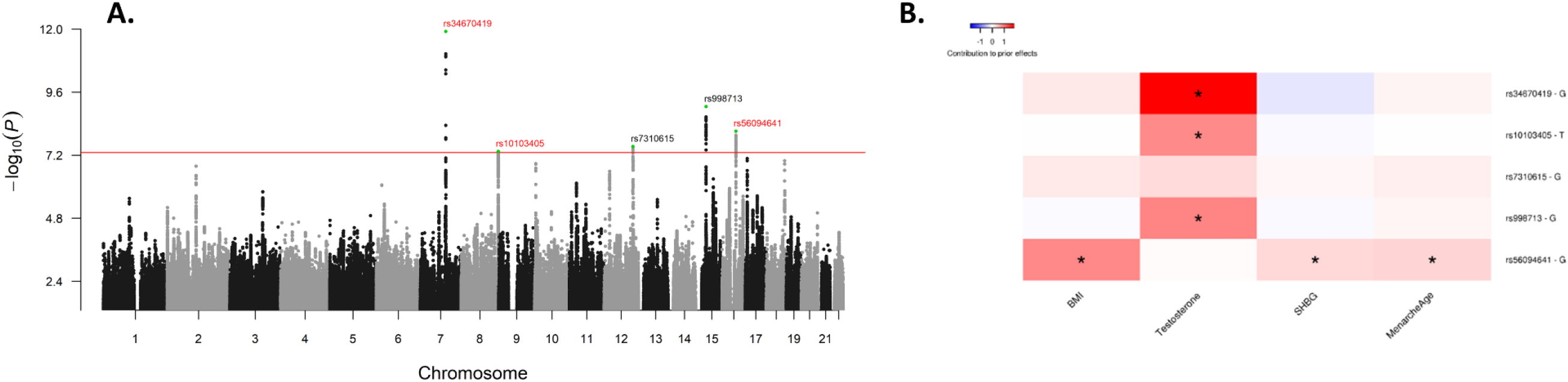
Manhattan plot of the –log_10_ permutation P values of the Bayesian Factor (P_BF_) of endometrial cancer risk (A) and heat map of prior contribution of SNPs of risk factors showing significant associations with endometrial cancer risk (B). Novel loci are annotated in red text and known risk loci in black. The red line indicates genome-wide significance at –log_10_(5 × 10^−8^). Asterisk in the heat map indicates a significant variant-trait association at –log_10_(5 × 10^−8^).

Using direct effects, we detected 13 endometrial cancer risk loci (P_d_ < 5 × 10^−8^), all of which have been reported previously by GWAS analyses (**Supplementary Figure S5**; **Supplementary Table S5**), suggesting that they either affect endometrial cancer risk directly or through other risk factors not included in the analysis. An imputed singleton variant at 7p14.3 (rs9639594) associated with endometrioid endometrial cancer risk based on direct effects. This variant was identified as a potential endometrioid endometrial cancer risk variant in a previous GWAS analysis; however, as previously described, its association with endometrial cancer needs to be further investigated due to the sparse LD at this region^5^. In our secondary posterior effects GWAS analysis, we identified 15 potential endometrial cancer risk loci (P_p_ < 5 × 10^−8^) (**Supplementary Figure S6A and C**; **Supplementary Table S6**). These loci had very large prior effects contributed by the risk factors included in the analysis (**Supplementary Figure S6B and D**), with all but one of them significantly associated with at least one risk factor.

### Replication of novel endometrial cancer susceptibility loci

We attempted to validate detected loci using a replication set consisting of three publicly available GWAS datasets (UK Biobank, FinnGen and the Japanese Biobank) and another GWAS dataset from the UK^26,27^. All three novel loci found in our primary multi-trait BF GWAS analysis displayed a concordant direction of effect in this independent replication set, and two loci (7q22.1 and 8q24.3) replicated with nominally significant associations with endometrial cancer risk observed (P < 0.05) (**Table 3**; **Supplementary Figure S7**). From the 15 potential loci identified from the secondary posterior effects analysis, 80% of these (12/15 loci) displayed a concordant direction of effect in the independent replication set. Meta-analysis of the replication set with the larger ECAC GWAS dataset improved the association of the three loci identified by multi-trait BF GWAS analysis with endometrial cancer risk, although the 16q12.2 locus showed some evidence of heterogeneity (**Supplementary Figure S7C**).

**Table 3.**
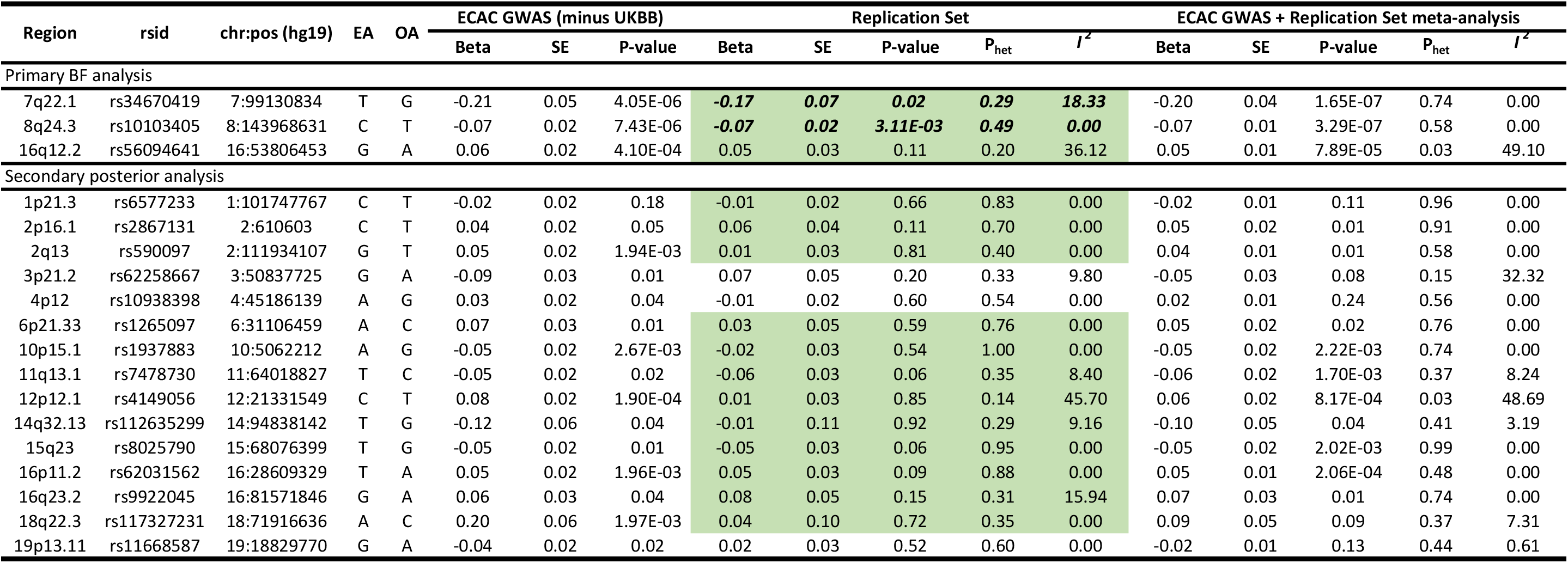
Replication of endometrial cancer risk variants identified by multivariate GWAS analyses. Abbreviations: EA - effect allele; OA - other allele; SE - standard error; P_het_ - heterogeneity p-value; *I* ^*2*^ - heterogeneity estimate; BF - Bayes factor; UKBB - UK Biobank Cells highligted in green are those which display a concordant direction of effect with the multivariate GWAS results in the replication set; variants with a nominal significance (P-value < 0.05) in the replication set are noted in bold italics

### Functional analyses of novel endometrial cancer susceptibility loci

We assessed quantitative trait loci (QTL) data from the Genotype-Tissue Expression (GTEx v8) project to determine whether candidate causal risk variants (determined using 100:1 log likelihood ratios; **Supplementary Table S7**) at the three novel loci identified by the primary BF analysis also represented associations with gene expression or splicing. At 7q22.1, candidate causal risk variants were associated with expression of *ZKSCAN5* (suprapubic skin) and *CYP3A7* (adrenal gland, visceral adipose and small intestine) (**Figure 4A-C**; **Supplementary Table S8**). Relevant to the association with testosterone levels at this locus, *CYP3A7* encodes an enzyme that metabolizes testosterone^35^ and we observed that the risk alleles associate with lower *CYP3A7* expression in the GTEx tissues. The lead expression QTL for *CYP3A7* in adrenal gland and visceral adipose (candidate causal variant rs45446698), is located only 128 bp from the summit of a *CYP3A7* promoter that has activity in adipocytes^36^ (promoter p1@CYP3A7; http://slidebase.binf.ku.dk/human_promoters/). *In silico* analysis predicted that this variant modifies several transcription factor binding motifs (**Supplementary Table S9**). These include motifs created by the risk allele that are bound by transcriptional repressors (MEIS1, FOXP1 and FOXD3) and whose expression is positively correlated with *CYP3A7* expression, in the adrenal gland or visceral adipose, providing a potential cis-regulatory mechanism for the QTL (**Supplementary Table S9**).

**Figure 4.**
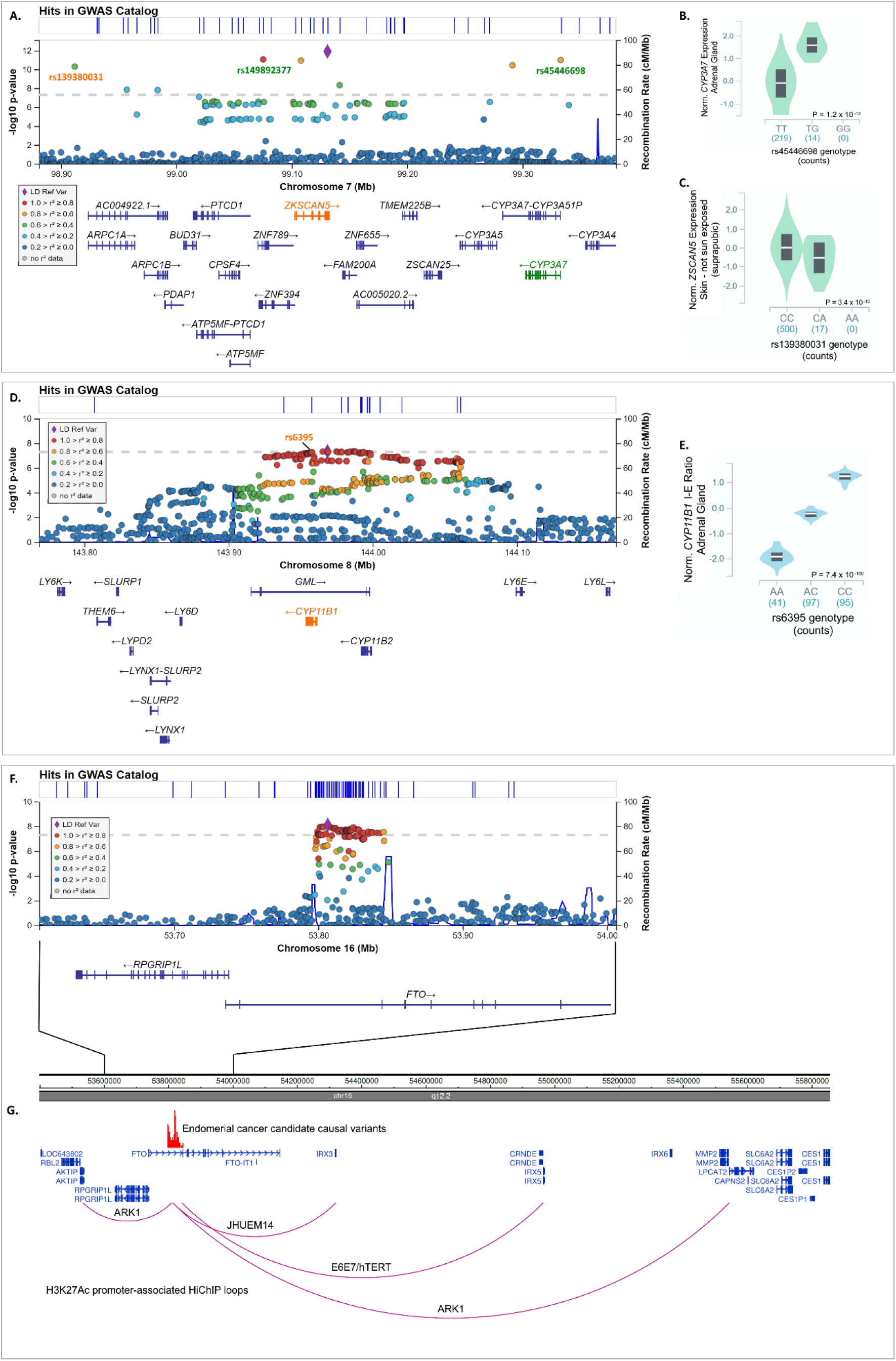
Novel endometrial cancer risk loci identified by multi-trait BF GWAS. Regional association plots for **A**. 7q22.1; **D**. 8q24.3 and; **F**. 16q12.22. Genetic variants at each locus are plotted by their genomic position (hg19) and multi-trait BF GWAS P-value –log_10_(P_BF_) for association with endometrial cancer risk on the left y-axis. Recombination rate (cM/Mb) is on the right y-axis and plotted as blue lines. The color of the circles indicates the level of linkage disequilibrium between each variant and the lead variant (purple diamond) from the 1000 Genomes 2014 EUR reference panel (see legend, inset). Expression and splicing quantitative trait loci variants (eQTL/sQTL) are labelled with associated genes highlighted in the same color. Violin plots of expression by genotype for eQTLs are provided for **B**. rs45446698 and *CYP3A7* (adrenal gland) and **C**. rs1139380031 and *ZKSCAN5* (skin not sun exposed – suprapubic). **E**. Violin plot of isoform-exon ratio (I-E Ratio) by genotype for sQTL rs6395 and *CYP11B1* (adrenal gland). **G**. Promoter-associated chromatin looping at 16q12.22 identified from HiChIP analysis of endometrial cancer cell lines. Promoter-associated loops that intersect with candidate causal variants (shown as red vertical lines) are shown as purple arcs.

None of the candidate causal risk variants at 8q24.3 represented associations with gene expression in GTEx tissues. However, eight of the candidate causal variants at this locus were joint lead splicing QTLs for *CYP11B1* in the adrenal gland (P = 7.40 × 10^−160^) (**Figure 4D and E**; **Supplementary Table S10**), with risk alleles associated with aberrant splicing. *CYP11B1* encodes an adrenal gland-specific enzyme that also metabolizes testosterone and 8q24.3 is another locus that is associated with testosterone levels.

We found no evidence that candidate causal risk variants at the 16q12.2 locus represented associations with gene expression or splicing in GTEx tissues. Therefore, we used available enhancer/promoter chromatin looping data from endometrial cell lines^32^ to identify five genes (*AKTIP, CRNDE, IRX3, IRX5* and *LPCAT2*) at this locus that are potentially targeted by risk variation through long-range interactions (**Figure 4F and G**). Notably, *IRX3* and *IRX5* have been established as causal genes for the association with BMI at this locus^37,38^.

## Discussion

Using genetic correlation, Mendelian randomization and multi-trait GWAS approaches we have identified novel endometrial cancer risk loci, and highlighted factors underlying endometrial cancer development. Genetic correlation analyses found a large number of traits to be significantly correlated with endometrial cancer predisposition, 24 of which remained significant after adjusting for the BMI. Mendelian randomization analyses found 90% of these to be likely causal for endometrial cancer development, although only the relationship for SHBG levels remained significant after adjusting for BMI. Using priors from multivariate Mendelian randomization analyses in a multi-trait BF GWAS approach, we detected three novel endometrial cancer risk loci, two of which replicated in an independent dataset. Secondary analysis using multi-trait posterior effects GWAS analysis highlighted another 15 potential endometrial cancer risk loci for further investigation.

Identifying genetically correlated traits can provide useful etiological insights into endometrial cancer and priorities likely causal relationships. Therefore, a fundamental goal of endometrial cancer epidemiology is to understand its relationships with other complex human phenotypes. One of our previous studies identified 14 non-cancer traits that were significantly correlated with endometrial cancer, all of which are either a proxy trait for obesity or are strongly and significantly genetically correlated with BMI^5^. By using the expanded number of traits in the UK Biobank and other consortia, we found 65 non-cancer phenotypes and 2 cancers (breast and ovarian cancers) that were genetically correlated with endometrial cancer. However, genetic correlation with endometrial cancer is subject to genetic confounding; for instance, traits might be genetically with endometrial cancer via their correlations with BMI. This was the case for about two thirds (43) of the 67 traits, as demonstrated by the observation that only 24 traits were still significantly genetically correlated with endometrial cancer after adjustment for BMI, most of which were related to body composition. These results support the previous assumption that although BMI is widely used as a proxy measure for obesity, the aspect of body composition most relevant for endometrial cancer risk is only partially captured by BMI^10^. In addition, genetic variation associated with traits not related to BMI, such as SHBG levels and relative age of first facial hair, were also shown to be significantly genetically correlated with endometrial cancer risk.

To date, Mendelian randomization approaches have identified several factors that affect endometrial cancer risk, including estradiol^13^, age at menarche^5,11^, HDL^14^, LDL^14^, testosterone^12^, and SHBG^12^. In this study, besides anthropometric traits, we also found that genetic propensity for non-anthropometric traits affected endometrial cancer risk, including SHBG and previously unreported traits such as basal metabolic rate and relative age of first facial hair. Propensity for increased SHBG levels and relative age of first facial hair provided protective effects on endometrial cancer risk, which was consistent with their negative genetic correlations with endometrial cancer.

Our primary multi-trait BF GWAS analysis identified three novel risk loci for endometrial cancer, two of which replicated in an independent dataset (7q22.1 and 8q24.3). Both of these novel endometrial cancer risk loci harbor genes encoding enzymes that metabolize testosterone (*CYP3A7* and *CYP11B1*). Estrogen, which plays a crucial role in endometrial carcinogenesis, is synthesized from testosterone; thus, perturbation of testosterone metabolic pathways is likely to affect endometrial cancer risk. It is expected decreased levels of the *CYP3A7* and *CYP11B1* transcripts would result in an increase of sex-hormone levels. Indeed, candidate causal variants at the 7q22.1 endometrial cancer risk signal associate with decreased expression of *CYP3A7*, increased total and bioavailable testosterone in women^12^, increased estradiol levels in men^12^, increased progesterone levels (in males and females combined)^39^ and increased bone mineral density^40^, a trait highly influenced by estrogen levels. Analyses at 7q22.1 highlighted candidate causal variant rs45446698 as a likely functional variant whose risk allele may reduce *CYP3A7* expression by generation of motifs bound by three transcriptional repressors. Consistent with this effect, two repressors (MEIS1 and FOXP1) also inhibit testosterone signaling through effects on the androgen receptor^41,42^. At the 8q24.3 locus, candidate causal endometrial cancer risk variants associate with aberrant splicing of *CYP11B1*, increased total and bioavailable testosterone in women^12^ and increased estrone levels in men^43^. Interestingly, associations with testosterone levels at both loci are highly female-specific with no significant association observed amongst men^12^.

The remaining novel risk locus identified by our multi-trait BF GWAS analysis is located at 16q12.2, intronic to the *FTO* gene. Although not statistically significant in the independent replication set (P > 0.05), the direction of effect appeared concordant. A highly pleiotropic locus, 16q12.2 was the first locus identified by GWAS of BMI and has subsequently been reported to associate with a large number of anthropometric and obesity-related traits, including waist and hip circumference, body fat percentage, visceral adipose levels, diabetes and metabolic syndrome (reviewed by Yang et al., 2017^44^). A recent GWAS reported that variants at this locus associate with breast cancer risk^45^. This is the first study to identify 16q12.2 as a potential endometrial cancer risk locus using GWAS analysis. The endometrial cancer risk signal is the same as that reported for BMI, with the endometrial cancer risk variant associating with increased BMI, as expected. Although obesity-related variants are located within the *FTO* gene, a range of experiments, including CRISPR gene editing, mouse models and chromatin capture assays, linked *IRX3* and *IRX5* to this phenotype^37,38^. Interrogation of promoter-associated loops in endometrial cell lines found interactions between endometrial cancer risk variants and *IRX3* and *IRX5*. We additionally identified the promoters of *AKTIP, CRNDE* and *LPCAT2* as interacting with endometrial cancer risk variation through chromatin looping at this locus, but their role, if any, in endometrial carcinogenesis is unclear.

The findings from the Mendelian randomization and GWAS analyses suggest therapeutic strategies to prevent or treat endometrial cancer (e.g. weight loss or reduction of testosterone levels). Weight loss, in particular that induced by bariatric surgery, has been shown to reduce endometrial cancer risk by up to 80% (reviewed by ^46^). However, there have been limited studies of the effect of weight loss on endometrial cancer survival and the few studies to date have been small and inconclusive^47^. Although there are a number of medications available to reduce testosterone levels, through a variety of mechanisms (reviewed in ^48^), there have also been very few studies that have intentionally targeted testosterone in endometrial cancer. Currently, it is not clear if testosterone exerts its effects on endometrial cancer susceptibility simply as a precursor to estrogen or through other pathways such as androgen receptor signaling. Intriguingly, an androgen receptor inhibitor that blocks testosterone signaling (enzalutamide) has been reported to inhibit proliferation of primary endometrial tumor cells^49^ and is currently being studied in combination with chemotherapy in a phase II trial of endometrial cancer (ClinicalTrials.gov Identifier: NCT02684227). Further studies are needed in this area to determine the molecular effects of testosterone on endometrial cancer development and its potential for therapeutic targeting.

This study demonstrates the strength of using the largest endometrial cancer GWAS dataset available and leveraging genetic information of risk factors to identify new susceptibility loci to endometrial cancer. The significance and directions of these new loci were also validated in an independent endometrial cancer GWAS dataset, though not all loci were confirmed. This indicates the necessity of a larger replication dataset to verify the findings in this study. All studies included in the discovery ECAC GWAS and the datasets included in the multi-trait bGWAS analysis were of European ancestry. Thus, we cannot say whether these variants are transferrable to other ethnic groups, a limitation of this study. The lack of availability of sex-stratified GWAS summary statistics for some traits may fail to detect some risk factors and consequently risk loci of endometrial cancer due to the existence of sex dimorphism for some traits^12,50-55^. As the UK Biobank, FinnGen, and Japanese Biobank did not include histology information of endometrial cancer cases, we were unable to validate the identified risk loci associated with endometrioid endometrial cancer. We were also unable to assess non-endometrioid endometrial cancer in our study. Due to the small number of cases in this subset (n = 1,230)^5^, we could not quantify the heritability of non-endometrioid endometrial and therefore could not run genetic correlation analyses.

In conclusion, we have used genetic approaches to comprehensively identify and assess risk factors for endometrial cancer. We were then able to leverage risk factor genetic information in a multi-trait GWAS analysis to identify novel endometrial cancer risk loci. We found the majority of risk factors for endometrial cancer are related to obesity and body composition. Multivariate Mendelian randomization analysis highlighted the importance of sex steroid hormones on endometrial cancer development, with three of the four independent risk factors identified related to sex hormone exposure (i.e. testosterone levels, SHBG levels and age at menarche). Indeed, two of the three novel loci identified by multi-trait GWAS analysis are strongly associated with testosterone levels in women. The findings of this study identify additional opportunities for hormone treatment and further support weight loss measures such as bariatric surgery to reduce the risk of developing endometrial cancer^56^.

## Data Availability

Summary-level GWAS results for endometrial cancer used that support the findings of this study are available from the NHGRI-EBI GWAS Catalog (https://www.ebi.ac.uk/gwas/downloads/summary-statistics), the Japanese Biobank (http://jenger.riken.jp/en/) and FinnGen (http://r2.finngen.fi/). The UK Biobank data used to perform GWAS of endometrial cancer risk was accessed via application number 25331. Summary-level GWAS results for all other traits used in this study can be downloaded from URLs provided in Supplementary Table S1. Other data generated or used during this study are included in this article and its supplementary information files or are available on reasonable request.

http://r2.finngen.fi/

http://jenger.riken.jp/en/

## Acknowledgements

This work was conducted using the UK Biobank Resource (application number 25331). We thank the participants and investigators of FinnGen study and the Biobank Japan Project. We would also like to acknowledge the Neale laboratory for generating the GWAS summary statistics of the UK Biobanks phenotypes and for making them publicly available. The Genotype-Tissue Expression (GTEx) Project was supported by the Common Fund of the Office of the Director of the National Institutes of Health, and by NCI, NHGRI, NHLBI, NIDA, NIMH, and NINDS. The data used for the analyses described in this manuscript were obtained from the GTEx Portal in February 2021. Data on lipid levels have been contributed by Global Lipids Genetics Consortium investigators and have been downloaded from http://csg.sph.umich.edu/willer/public/lipids2013/. Data on glycemic traits have been contributed by MAGIC investigators and have been downloaded from www.magicinvestigators.org. The breast cancer genome-wide association analyses were supported by the Government of Canada through Genome Canada and the Canadian Institutes of Health Research, the ‘Ministère de l’Économie, de la Science et de l’Innovation du Québec’ through Genome Québec and grant PSR-SIIRI-701, The National Institutes of Health (U19 CA148065, X01HG007492), Cancer Research UK (C1287/A10118, C1287/A16563, C1287/A10710) and The European Union (HEALTH-F2-2009-223175 and H2020 633784 and 634935). All studies and funders are listed in Michailidou et al (Nature, 2017). The ovarian cancer genome-wide association analyses were supported by the U.S. National Institutes of Health (CA1X01HG007491-01 (C.I. Amos), U19-CA148112 (T.A. Sellers), R01-CA149429 (C.M. Phelan) R01-CA058598 (M.T. Goodman) and R01-CA211707, R01-CA204954, and R01-CA211575 (S.A. Gayther); Canadian Institutes of Health Research (MOP-86727 (L.E. Kelemen)) and the Ovarian Cancer Research Fund (A. Berchuck). The COGS project was funded through a European Commission’s Seventh Framework Programme grant (agreement number 223175 – HEALTH-F2–2009-223175). The Gynaecological Oncology Biobank at Westmead, a member of the Australasian Biospecimen Network-Oncology group, was funded by the National Health and Medical Research Council Enabling Grants ID 310670 & ID 628903 and the Cancer Institute NSW Grants ID 12/RIG/1-17 & 15/RIG/1-16. All studies and funders are listed in Phelan et al (2017).

We thank the many individuals who participated in the Endometrial Cancer Association Consortium and the numerous institutions and their staff who supported recruitment. We particularly thank the efforts of Deborah Thompson. The endometrial cancer genome-wide association analyses were supported by the National Health and Medical Research Council of Australia (APP552402, APP1031333, APP1109286, APP1111246 and APP1061779), the U.S. National Institutes of Health (R01-CA134958), European Research Council (EU FP7 Grant), Wellcome Trust Centre for Human Genetics (090532/Z/09Z) and Cancer Research UK. OncoArray genotyping of ECAC cases was performed with the generous assistance of the Ovarian Cancer Association Consortium (OCAC), which was funded through grants from the U.S. National Institutes of Health (CA1X01HG007491-01 (C.I. Amos), U19-CA148112 (T.A. Sellers), R01-CA149429 (C.M. Phelan) and R01-CA058598 (M.T. Goodman); Canadian Institutes of Health Research (MOP-86727 (L.E. Kelemen)) and the Ovarian Cancer Research Fund (A. Berchuck). We particularly thank the efforts of Cathy Phelan. OncoArray genotyping of the BCAC controls was funded by Genome Canada Grant GPH-129344, NIH Grant U19 CA148065, and Cancer UK Grant C1287/A16563. All studies and funders are listed in O’Mara et al (2018). Full acknowledgements and funding for ECAC can be found in the Supplementary Note.

## Supplementary data

Supplementary data includes seven supplementary figures. A full list of Endometrial Cancer Association Consortium collaborators and their affiliations, together with Endometrial Cancer Association Consortium Acknowledgements, are also presented in Supplementary data.

Supplementary Tables are provided in a separate excel file.

## References

1. Bray, F. et al. Global cancer statistics 2018: GLOBOCAN estimates of incidence and mortality worldwide for 36 cancers in 185 countries. CA Cancer J Clin 68, 394–424 (2018).

2. Lortet-Tieulent, J., Ferlay, J., Bray, F. & Jemal, A. International Patterns and Trends in Endometrial Cancer Incidence, 1978-2013. Jnci-Journal of the National Cancer Institute 110, 354–361 (2018).

3. Siegel, R.L., Miller, K.D., Fuchs, H.E. & Jemal, A. Cancer Statistics, 2021. CA Cancer J Clin 71, 7–33 (2021).

4. Zhang, S. et al. Cancer incidence and mortality in China, 2015. Journal of the National Cancer Center 1, 2–11 (2021).

5. O’Mara, T.A. et al. Identification of nine new susceptibility loci for endometrial cancer. Nature Communications 9(2018).

6. Spurdle, A.B. et al. Genome-wide association study identifies a common variant associated with risk of endometrial cancer. Nature Genetics 43, 451-+ (2011).

7. Cheng, T.H.T. et al. Five endometrial cancer risk loci identified through genome-wide association analysis. Nature Genetics 48, 667–674 (2016).

8. Setiawan, V.W. et al. Type I and II endometrial cancers: have they different risk factors? Journal of Clinical Oncology 31, 2607–2618 (2013).

9. Nead, K.T. et al. Evidence of a causal association between insulinemia and endometrial cancer: a Mendelian randomization analysis. JNCI: Journal of the National Cancer Institute 107(2015).

10. Painter, J.N. et al. Genetic Risk Score Mendelian Randomization Shows that Obesity Measured as Body Mass Index, but not Waist: Hip Ratio, Is Causal for Endometrial Cancer. Cancer Epidemiology Biomarkers & Prevention 25, 1503–1510 (2016).

11. Day, F.R. et al. Genomic analyses identify hundreds of variants associated with age at menarche and support a role for puberty timing in cancer risk. Nature Genetics 49, 834–841 (2017).

12. Ruth, K.S. et al. Using human genetics to understand the disease impacts of testosterone in men and women. Nature Medicine 26, 252–258 (2020).

13. Thompson, D.J. et al. CYP19A1 fine-mapping and Mendelian randomization: estradiol is causal for endometrial cancer. Endocrine-Related Cancer 23, 77–91 (2016).

14. Kho, P.F. et al. Mendelian randomization analyses suggest a role for cholesterol in the development of endometrial cancer. Int J Cancer (2020).

15. Mounier, N. & Kutalik, Z. bGWAS: an R package to perform Bayesian Genome Wide Association Studies. Bioinformatics (2020).

16. McDaid, A.F. et al. Bayesian association scan reveals loci associated with human lifespan and linked biomarkers. (2017).

17. Timmers, P.R. et al. Genomics of 1 million parent lifespans implicates novel pathways and common diseases and distinguishes survival chances. eLife 8, 1–40 (2019).

18. Ghodsian, N. et al. Electronic Health Record-Based Genome-Wide Meta-Analysis and Mendelian Randomization Identify Metabolic and Phenotypic Consequences of Non-Alcoholic Fatty Liver Disease. preprint (2020).

19. Sudlow, C. et al. UK biobank: an open access resource for identifying the causes of a wide range of complex diseases of middle and old age. PLoS Med 12, e1001779 (2015).

20. Bulik-Sullivan, B. et al. LD score regression distinguishes confounding from polygenicity in genome-wide association studies. Nature Genetics 47, 291–295 (2015).

21. Zhu, Z. et al. Causal associations between risk factors and common diseases inferred from GWAS summary data. Nature Communications 9(2018).

22. Yang, J., Lee, S.H., Goddard, M.E. & Visscher, P.M. GCTA: a tool for genome-wide complex trait analysis. Am J Hum Genet 88, 76–82 (2011).

23. Yang, J. et al. Conditional and joint multiple-SNP analysis of GWAS summary statistics identifies additional variants influencing complex traits. Nat Genet 44, 369-375, S361-363 (2012).

24. Hemani, G. et al. The MR-Base platform supports systematic causal inference across the human phenome. Elife 7(2018).

25. Ishigaki, K. et al. Large-scale genome-wide association study in a Japanese population identifies novel susceptibility loci across different diseases. Nature Genetics 4(2020).

26. Evans, D.G.R. et al. Breast cancer pathology and stage are better predicted by risk stratification models that include mammographic density and common genetic variants. Breast cancer research and treatment 176, 141–148 (2019).

27. Ryan, N.A.J. et al. The proportion of endometrial tumours associated with Lynch syndrome (PETALS): A prospective cross-sectional study. PLoS Med 17, e1003263 (2020).

28. Mbatchou, J. et al. Computationally efficient whole genome regression for quantitative and binary traits. Nature Genetics (2021).

29. Willer, C.J., Li, Y. & Abecasis, G.R. METAL: fast and efficient meta-analysis of genomewide association scans. Bioinformatics 26, 2190–2191 (2010).

30. Munz, M. et al. Qtlizer: comprehensive QTL annotation of GWAS results. Sci Rep 10, 20417 (2020).

31. Consortium, T.G. The GTEx Consortium atlas of genetic regulatory effects across human tissues. Science 369, 1318–1330 (2020).

32. O’Mara, T.A., Spurdle, A.B., Glubb, D.M. & Consortium, E.C.A. Analysis of Promoter-Associated Chromatin Interactions Reveals Biologically Relevant Candidate Target Genes at Endometrial Cancer Risk Loci. Cancers 11(2019).

33. Bailey, T.L. et al. MEME SUITE: tools for motif discovery and searching. Nucleic Acids Res 37, W202–208 (2009).

34. Tang, Z., Kang, B., Li, C., Chen, T. & Zhang, Z. GEPIA2: an enhanced web server for large-scale expression profiling and interactive analysis. Nucleic Acids Res 47, W556–W560 (2019).

35. Kandel, S.E., Han, L.W., Mao, Q.C. & Lampe, J.N. Digging Deeper into CYP3A Testosterone Metabolism: Kinetic, Regioselectivity, and Stereoselectivity Differences between CYP3A4/5 and CYP3A7. Drug Metabolism and Disposition 45, 1266–1275 (2017).

36. Clst, F.C.a.t.R.P.a. et al. A promoter-level mammalian expression atlas. Nature 507, 462–470 (2014).

37. Claussnitzer, M. et al. FTO Obesity Variant Circuitry and Adipocyte Browning in Humans. N Engl J Med 373, 895–907 (2015).

38. Smemo, S. et al. Obesity-associated variants within FTO form long-range functional connections with IRX3. Nature 507, 371-+ (2014).

39. Ruth, K.S. et al. Genome-wide association study with 1000 genomes imputation identifies signals for nine sex hormone-related phenotypes. European Journal of Human Genetics 24, 284–290 (2016).

40. Morris, J.A. et al. An atlas of genetic influences on osteoporosis in humans and mice. Nature Genetics 51, 258–266 (2019).

41. Takayama, K. et al. FOXP1 is an androgen-responsive transcription factor that negatively regulates androgen receptor signaling in prostate cancer cells. Biochemical and Biophysical Research Communications 374, 388–393 (2008).

42. Cui, L. et al. MEIS1 functions as a potential AR negative regulator. Experimental Cell Research 328, 58–68 (2014).

43. Eriksson, A.L. et al. Genetic Determinants of Circulating Estrogen Levels and Evidence of a Causal Effect of Estradiol on Bone Density in Men. J Clin Endocrinol Metab 103, 991–1004 (2018).

44. Yang, Q.Y., Xiao, T.C., Guo, J. & Su, Z.Q. Complex Relationship between Obesity and the Fat Mass and Obesity Locus. International Journal of Biological Sciences 13, 615–629 (2017).

45. Michailidou, K. et al. Association analysis identifies 65 new breast cancer risk loci. Nature 551, 92–94 (2017).

46. Njoku, K., Abiola, J., Russell, J. & Crosbie, E.J. Endometrial cancer prevention in high-risk women. Best Pract Res Clin Obstet Gynaecol 65, 66–78 (2020).

47. Kitson, S. et al. Interventions for weight reduction in obesity to improve survival in women with endometrial cancer. Cochrane Database Syst Rev 2, CD012513 (2018).

48. Crawford, E.D. et al. Androgen-targeted therapy in men with prostate cancer: evolving practice and future considerations. Prostate Cancer Prostatic Dis 22, 24–38 (2019).

49. Tangen, I.L. et al. Androgen receptor as potential therapeutic target in metastatic endometrial cancer. Oncotarget 7, 49289–49298 (2016).

50. Flynn, E. et al. Sex-specific genetic effects across biomarkers. European Journal of Human Genetics 29, 154–163 (2021).

51. Graham, S.E. et al. Sex-specific and pleiotropic effects underlying kidney function identified from GWAS meta-analysis. Nature Communications 10(2019).

52. Khramtsova, E.A. et al. Sex differences in the genetic architecture of obsessive– compulsive disorder. American Journal of Medical Genetics, Part B: Neuropsychiatric Genetics 180, 351–364 (2019).

53. Link, J.C. & Reue, K. Genetic Basis for Sex Differences in Obesity and Lipid Metabolism. Annu Rev Nutr 37, 225–245 (2017).

54. Randall, J.C. et al. Sex-stratified Genome-wide Association Studies Including 270,000 Individuals Show Sexual Dimorphism in Genetic Loci for Anthropometric Traits. PLoS Genetics 9(2013).

55. Lagou, V. et al. Sex-dimorphic genetic effects and novel loci for fasting glucose and insulin variability. Nat Commun 12, 24 (2021).

56. MacKintosh, M.L. & Crosbie, E.J. Prevention Strategies in Endometrial Carcinoma. Curr Oncol Rep 20, 101 (2018).

