## Supplementary Information and Figures for "Multi-trait genome-wide association study identifies novel endometrial cancer risk loci that are associated with obesity or female testosterone levels"

**Supplementary Tables (in excel document)**

**Supplementary Table S1.** Phenotypes assessed and download information.

**Supplementary Table S2.** Correlation between prior and observed effects of endometrial cancer risk between different p-value thresholds for selection of instrumental variables included in Mendelian Randomisation.

**Supplementary Table S3.** Genetic correlations between phenotypes and endometrial cancer with or without adjustment for BMI.

**Supplementary Table S4.** Mendelian randomisation results with endometrial cancer risk for traits significantly correlated with endometrial cancer risk after adjustment for BMI. Relationships between traits and endometrial cancer risk were additionally assessed adjusting for BMI.

**Supplementary Table S5.** Genome-wide significant variant associations with risk of endometrial cancer based on direct effects by bGWAS.

**Supplementary Table S6.** Novel genome-wide significant variant associations with risk of endometrial cancer based on posterior effects by bGWAS.

**Supplementary Table S7.** Candidate causal risk variants at the three novel loci identified by the primary Bayes factor GWAS analysis.

**Supplementary Table S8.** Candidate causal endometrial cancer risk variant eQTLs.

**Supplementary Table S9.** Eight lead sQTLs for CYP11B1 in the adrenal gland.

**Supplementary Table S10.** MaxEntScan results of candidate causal risk variants of CYP11B1.

**Supplementary Figures**

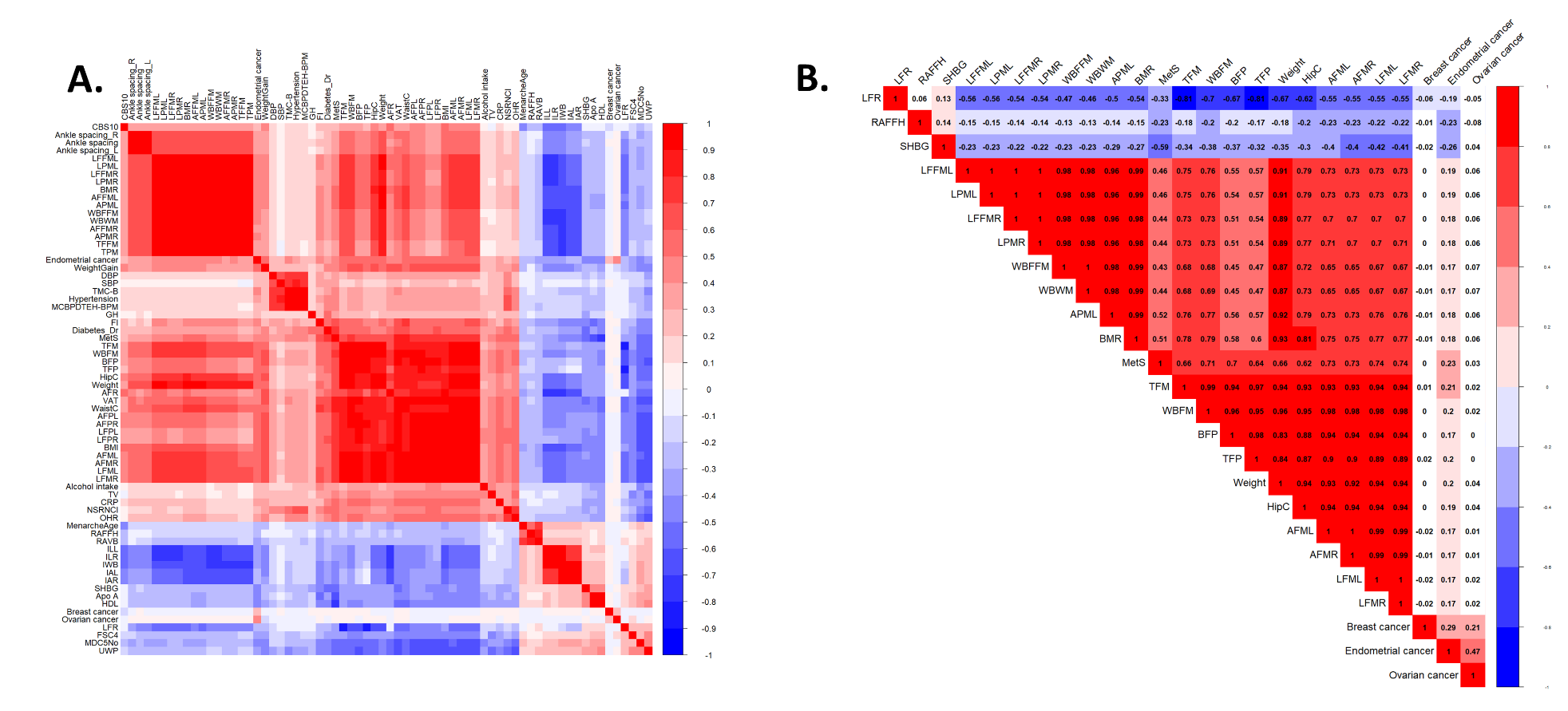

**Supplementary Figure S1. Heatmap of genetic correlations between endometrial cancer risk and (A) the 67 traits at FDR < 0.01, and (B) the 24 traits that remained significantly correlated with endometrial cancer after adjustment for BMI.** Abbreviations can be found in Supplementary Table S2.

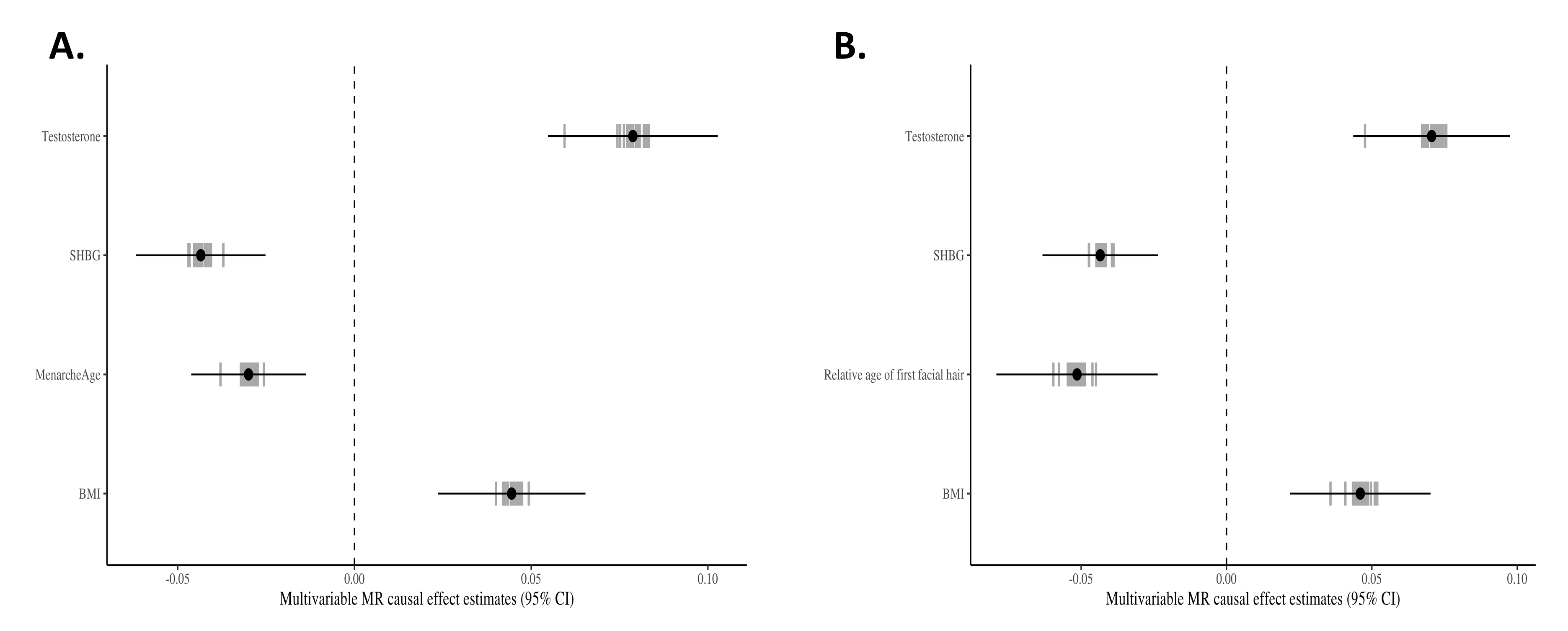

**Supplementary Figure S2. Coefficients plot for the four risk factors used to create the prior in multi-trait GWAS analysis of the risk of (A) endometrial cancer all histologies and (B) endometrioid histology.** The black dot is the multivariable causal effect estimate and the horizontal line the 95% interval from the multivariable Mendelian randomisation model using all chromosomes for each risk factor; whereas grey vertical bars represents the 22 per-chromosome estimates.

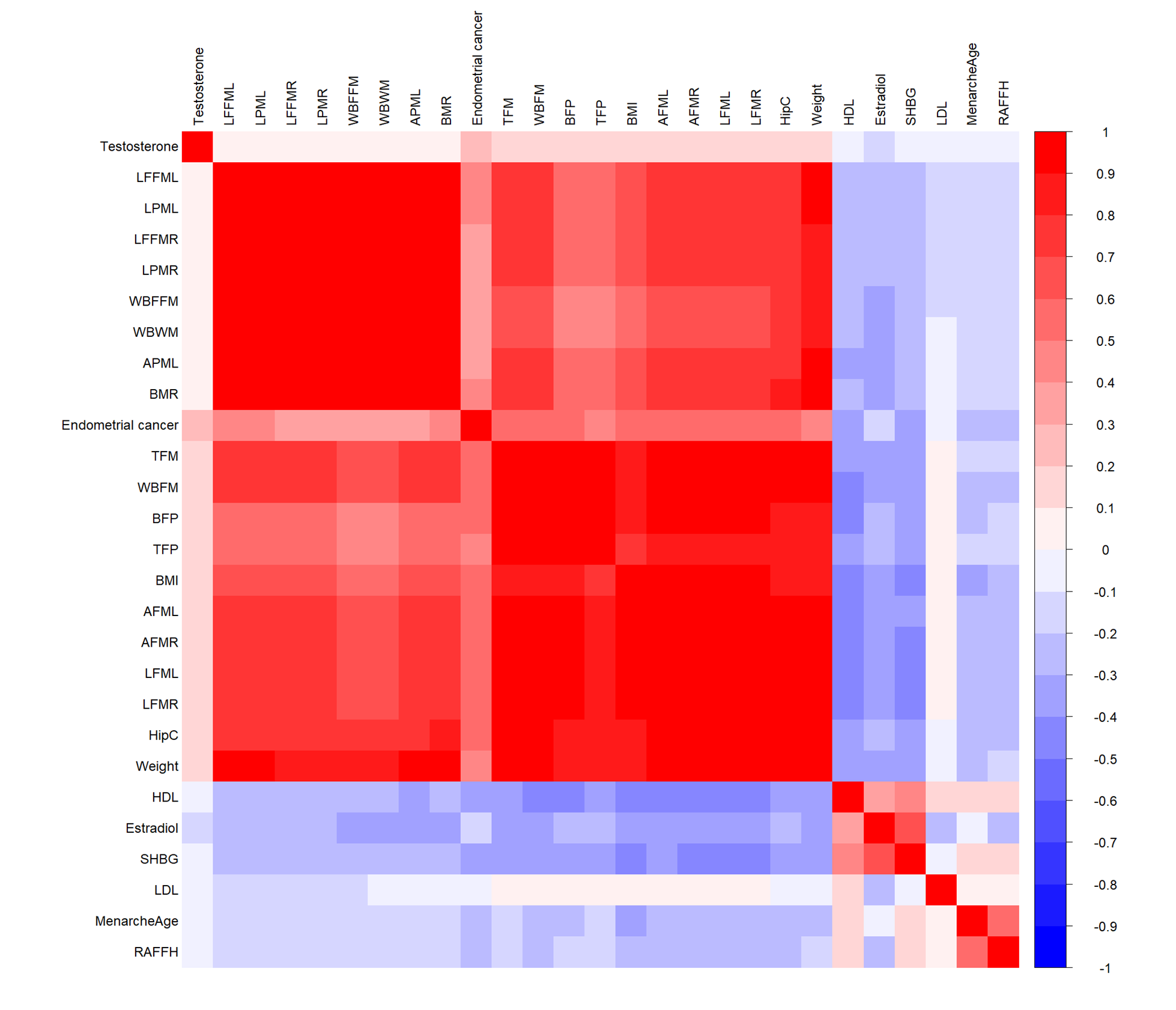

**Supplementary Figure S3. Heatmap of pairwise correlations of the 27 traits that were included in the multi-trait GWAS analysis of endometrial cancer risk.** Abbreviations can be found in Supplementary Table S2.

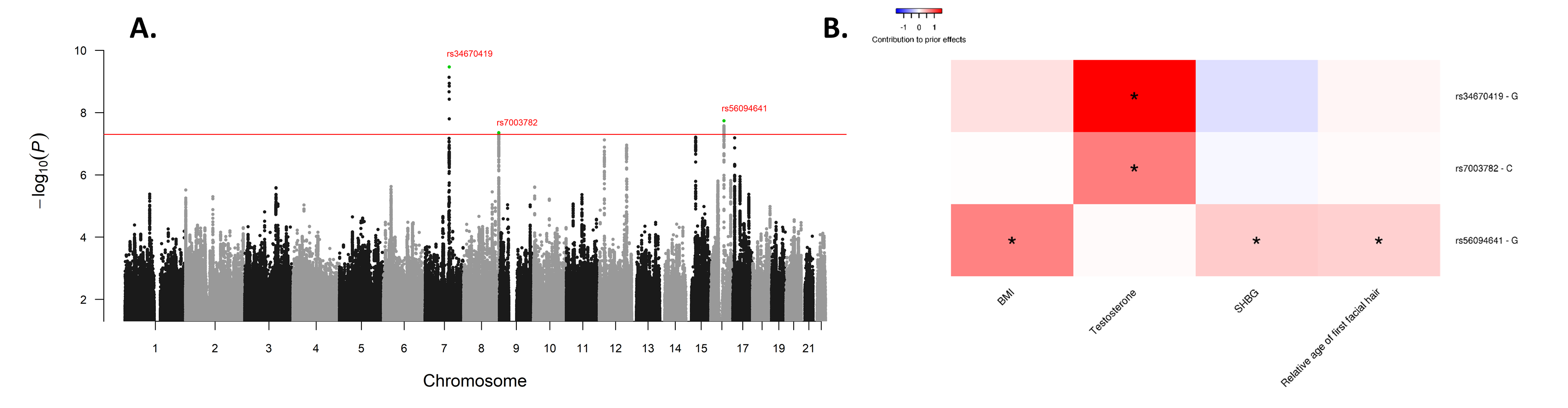

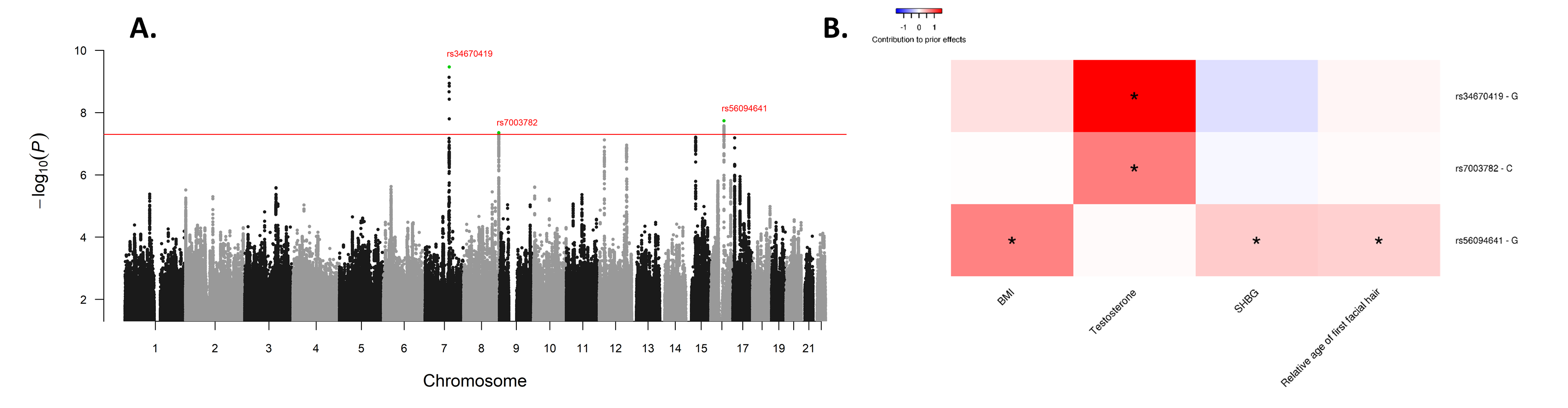

**Supplementary Figure S4. Manhattan plot of the -log_10_ permutation P values of the Bayesian factor (P_BF_) of endometrioid endometrial cancer risk (A) and heatmap of prior contribution of SNPs of risk factors showing significant association with endometrioid endometrial cancer risk (B).** Potentially novel loci were annotated. The red line indicates the genome-wide significance at -log_10_(5 × 10^-8^). Asterisk in the heatmap indicates a significant variant-trait association at -log_10_(5 × 10^-8^).

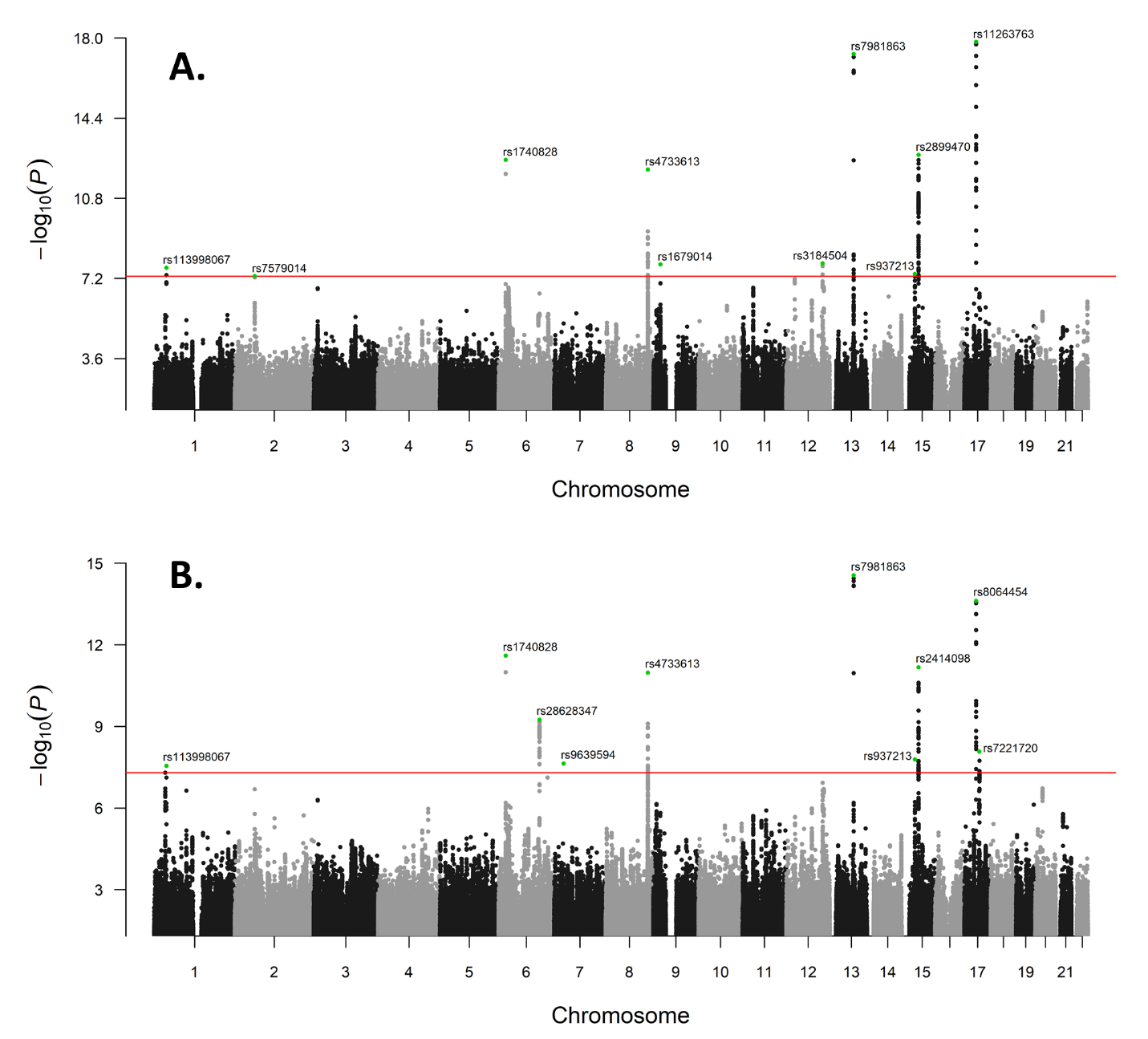

**Supplementary Figure S5. Manhattan plot of the -log_10_ permutation P values of the direct effects (P_d_) of the risks of (A) endometrial cancer all histologies and (B) endometrioid endometrial cancer.** SNPs displayed were independent variants achieving genome-wide significance (GWS), all of which were known risk variants. The red line indicates the GWS at –log_10_(5 × 10^-8^).

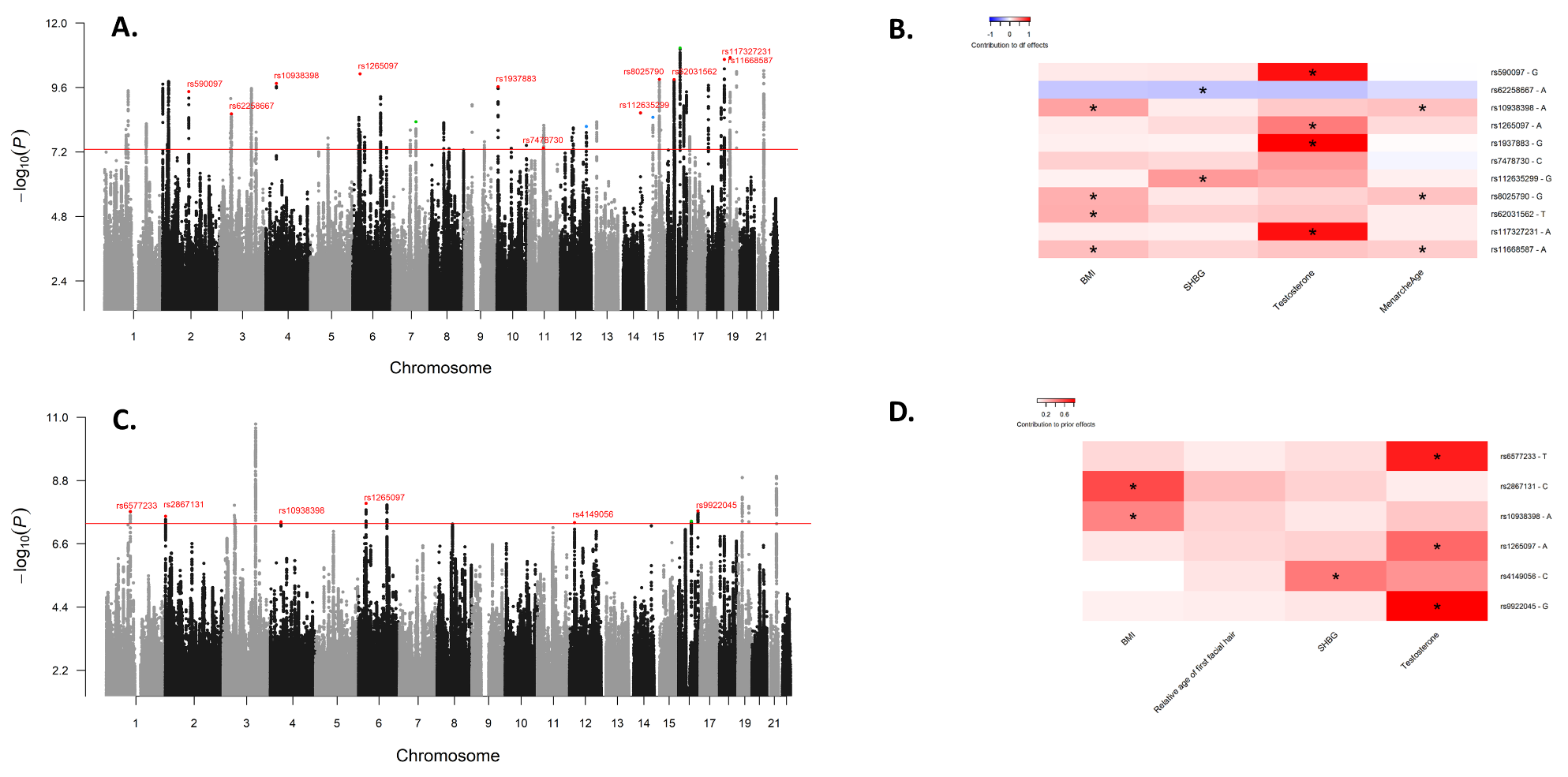

**Supplementary Figure S6. Manhattan plot of the -log_10_ permutation P values of the posterior effect (P_p_) of endometrial cancer risk (A. all histologies; C. endometrioid histology) and heatmap of prior contribution of SNPs of risk factors showing significant associations with endometrial cancer risk (B. all histologies; D. endometrioid histology).** Lead SNPs, which were nominally significant (P<0.05) in the O’Mara *et al* 2018 endometrial cancer GWAS^3^ but reached genome-wide significance in the posterior effect in this study are highlighted; two known SNPs are highlighted in blue and two SNPs detected by Bayes factors in green, whereas potential novel SNPs are highlighted and annotated in red. The red line indicates the genome-wide significance at -log_10_(5 × 10^-8^). Asterisk in the heatmap indicates a significant variant-trait association at -log_10_(5 × 10^-8^).

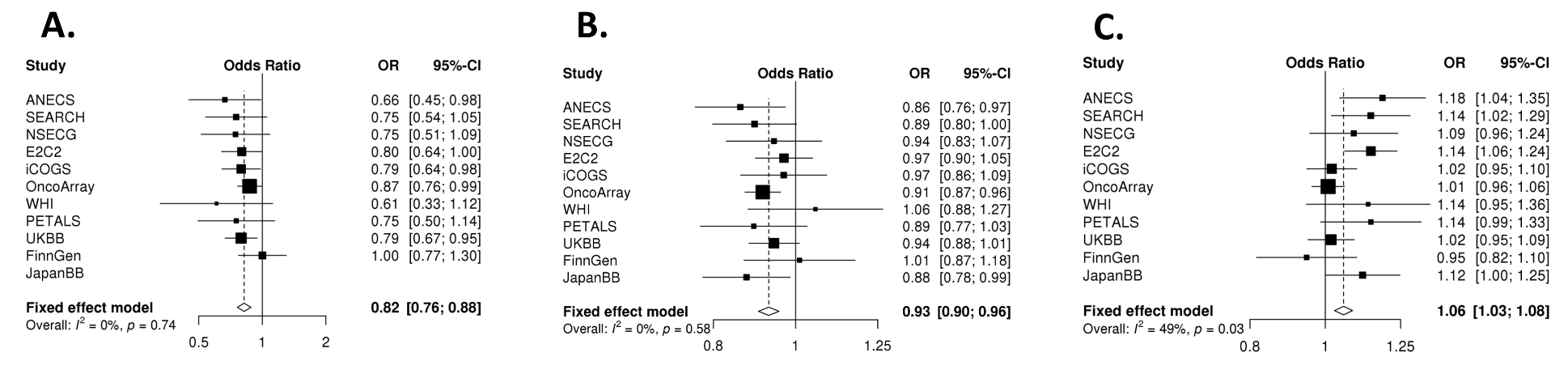

**Supplementary Figure S7. Forest plots for the replication meta-analysis of the lead variants at the three novel loci identified by multi-trait Bayes factor GWAS. A.** 7q22.1 rs34670419; **B.** 8q24.3 rs10103405; **C.** 16q12.22 rs56094641.

**Endometrial Cancer Association Consortium Collaborators**

Frederic Amant^1^, Daniela Annibali^1^, Katie Ashton^2-4^, John Attia^2, 5^, Paul L. Auer^6, 7^, Matthias W. Beckmann^8^, Amanda Black^9^, Louise Brinton^9^, Daniel D. Buchanan^10-13^, Stephen J. Chanock^14^, Chu Chen^15^, Maxine M. Chen^16^, Timothy H.T. Cheng^17^, Linda S. Cook^18, 19^, Marta Crous-Bous^16, 20^, Kamila Czene^21^, Immaculata De Vivo^16, 20^, Joe Dennis^22^, Thilo Dörk^23^, Sean C. Dowdy^24^, Alison M. Dunning^25^, Matthias Dürst^26^, Douglas F. Easton^22, 25^, Arif B. Ekici^27^, Peter A. Fasching^8, 28^, Brooke L. Fridley^29^, Christine M. Friedenreich^19^, Montserrat García-Closas^14^, Mia M. Gaudet^30^, Graham G. Giles^11, 31, 32^, Dylan M. Glubb^33^, Ellen L. Goode^34^, Christopher A. Haiman^35^, Per Hall^21, 36^, Susan E. Hankinson^20, 37^, Catherine S. Healey^25^, Alexander Hein^8^, Peter Hillemanns^23^, Shirley Hodgson^38^, Erling Hoivik^39, 40^, Elizabeth G. Holliday^2, 5^, David J. Hunter^16, 41^, Angela Jones^17^, Peter Kraft^16, 42^, Camilla Krakstad^39, 40^, Diether Lambrechts^43, 44^, Loic Le Marchand^45^, Xiaolin Liang^46^, Annika Lindblom^47, 48^, Jolanta Lissowska^49^, Jirong Long^50^, Lingeng Lu^51^, Anthony M. Magliocco^52^, Lynn Martin^53^, Mark McEvoy^5^, Roger L. Milne^11, 31, 32^, Miriam Mints^54^, Rami Nassir^55^, Tracy A. O'Mara^33^, Irene Orlow^46^, Geoffrey Otton^56^, Claire Palles^17^, Paul D.P. Pharoah^22, 25^, Loreall Pooler^35^, Tony Proietto^56^, Timothy R. Rebbeck^57, 58^, Stefan P. Renner^59^, Harvey A. Risch^51^, Matthias Rübner^59^, Ingo Runnebaum^26^, Carlotta Sacerdote^60, 61^, Gloria E. Sarto^62^, Fredrick Schumacher^63^, Rodney J. Scott^2, 4, 64^, V. Wendy Setiawan^35^, Mitul Shah^25^, Xin Sheng^35^, Xiao-Ou Shu^50^, Melissa C. Southey^10, 31, 32^, Amanda B. Spurdle^33^, Emma Tham^47, 65^, Deborah J. Thompson^22^, Ian Tomlinson^17, 53^, Jone Trovik^39, 40^, Constance Turman^16^, David Van Den Berg^35^, Zhaoming Wang^9^, Penelope M. Webb^66^, Nicolas Wentzensen^9^, Stacey J. Winham^67^, Lucy Xia^35^, Yong-Bing Xiang^68^, Hannah P. Yang^9^, Herbert Yu^45^, Wei Zheng^50^

^1^ Department of Obstetrics and Gynecology, Division of Gynecologic Oncology, University Hospitals KU Leuven, University of Leuven, Leuven, Belgium. ^2^ Hunter Medical Research Institute, John Hunter Hospital, Newcastle, New South Wales, Australia. ^3^ Centre for Information Based Medicine, University of Newcastle, Callaghan, New South Wales, Australia.
^4^ Discipline of Medical Genetics, School of Biomedical Sciences and Pharmacy, Faculty of Health, University of Newcastle, Callaghan, New South Wales, Australia. ^5^ Centre for Clinical Epidemiology and Biostatistics, School of Medicine and Public Health, University of Newcastle, Callaghan, New South Wales, Australia. ^6^ Cancer Prevention Program, Fred Hutchinson Cancer Research Center, Seattle, WA, USA. ^7^ Zilber School of Public Health, University of Wisconsin-Milwaukee, Milwaukee, WI, USA. ^8^ Department of Gynecology and Obstetrics, Comprehensive Cancer Center ER-EMN, University Hospital Erlangen, Friedrich-Alexander-University Erlangen-Nuremberg, Erlangen, Germany. ^9^ Division of Cancer Epidemiology and Genetics, National Cancer Institute, Bethesda, MD, USA. ^10^ Department of Clinical Pathology, The University of Melbourne, Melbourne, Victoria, Australia. ^11^ Centre for Epidemiology and Biostatistics, Melbourne School of Population and Global Health, The University of Melbourne, Melbourne, Victoria, Australia. ^12^ Genomic Medicine and Family Cancer Clinic, Royal Melbourne Hospital, Parkville, Victoria, Australia. ^13^ University of Melbourne Centre for Cancer Research, Victorian Comprehensive Cancer Centre, Parkville, Victoria, Australia. ^14^ Division of Cancer Epidemiology and Genetics, National Cancer Institute, National Institutes of Health, Department of Health and Human Services, Bethesda, MD, USA. ^15^ Epidemiology Program, Fred Hutchinson Cancer Research Center, Seattle, WA, USA. ^16^ Department of Epidemiology, Harvard T.H. Chan School of Public Health, Boston, MA, USA. ^17^ Wellcome Trust Centre for Human Genetics and Oxford NIHR Biomedical Research Centre, University of Oxford, Oxford, UK. ^18^ University of New Mexico Health Sciences Center, University of New Mexico, Albuquerque, NM, USA. ^19^ Department of Cancer Epidemiology and Prevention Research, Alberta Health Services, Calgary, AB, Canada. ^20^ Channing Division of Network Medicine, Department of Medicine, Brigham and Women's Hospital and Harvard Medical School, Boston, MA, USA. ^21^ Department of Medical Epidemiology and Biostatistics, Karolinska Institutet, Stockholm, Sweden. ^22^ Centre for Cancer Genetic Epidemiology, Department of Public Health and Primary Care, University of Cambridge, Cambridge, UK. ^23^ Gynaecology Research Unit, Hannover Medical School, Hannover, Germany. ^24^ Department of Obstetrics and Gynecology, Division of Gynecologic Oncology, Mayo Clinic, Rochester, MN, USA. ^25^ Centre for Cancer Genetic Epidemiology, Department of Oncology, University of Cambridge, Cambridge, UK.
^26^ Department of Gynaecology, Jena University Hospital - Friedrich Schiller University, Jena, Germany. ^27^ Institute of Human Genetics, University Hospital Erlangen, Friedrich-Alexander University Erlangen-Nuremberg, Comprehensive Cancer Center Erlangen-EMN, Erlangen, Germany. ^28^ David Geffen School of Medicine, Department of Medicine Division of Hematology and Oncology, University of California at Los Angeles, Los Angeles, CA, USA.
^29^ Department of Biostatistics, Kansas University Medical Center, Kansas City, KS, USA.
^30^ Department of Population Science, American Cancer Society, Atlanta, GA, USA. ^31^ Cancer Epidemiology Division, Cancer Council Victoria, Melbourne, Victoria, Australia. ^32^ Precision Medicine, School of Clinical Sciences at Monash Health, Monash University, Clayton, Victoria, Australia. ^33^ Department of Genetics and Computational Biology, QIMR Berghofer Medical Research Institute, Brisbane, Queensland, Australia. ^34^ Department of Health Science Research, Division of Epidemiology, Mayo Clinic, Rochester, MN, USA. ^35^ Department of Preventive Medicine, Keck School of Medicine, University of Southern California, Los Angeles, CA, USA. ^36^ Department of Oncology, Södersjukhuset, Stockholm, Sweden. ^37^ Department of Biostatistics & Epidemiology, University of Massachusetts, Amherst, Amherst, MA, USA. ^38^ Department of Clinical Genetics, St George's, University of London, London, UK. ^39^ Centre for Cancer Biomarkers CCBIO, Department of Clinical Science, University of Bergen, Bergen, Norway.
^40^ Department of Obstetrics and Gynecology, Haukeland University Hospital, Bergen, Norway. ^41^ Nuffield Department of Population Health, University of Oxford, Oxford, UK. ^42^ Program in Genetic Epidemiology and Statistical Genetics, Harvard T.H. Chan School of Public Health, Boston, MA, USA. ^43^ VIB Center for Cancer Biology, Leuven, Belgium. ^44^ Laboratory for Translational Genetics, Department of Human Genetics, University of Leuven, Leuven, Belgium. ^45^ Epidemiology Program, University of Hawaii Cancer Center, Honolulu, HI, USA.
^46^ Department of Epidemiology and Biostatistics, Memorial Sloan-Kettering Cancer Center, New York, NY, USA. ^47^ Department of Molecular Medicine and Surgery, Karolinska Institutet, Stockholm, Sweden. ^48^ Department of Clinical Genetics, Karolinska University Hospital, Stockholm, Sweden. ^49^ Department of Cancer Epidemiology and Prevention, M. Sklodowska-Curie Cancer Center, Oncology Institute, Warsaw, Poland. ^50^ Division of Epidemiology, Department of Medicine, Vanderbilt Epidemiology Center, Vanderbilt-Ingram Cancer Center, Vanderbilt University School of Medicine, Nashville, TN, USA. ^51^ Chronic Disease Epidemiology, Yale School of Public Health, New Haven, CT, USA. ^52^ Department of Anatomic Pathology, Moffitt Cancer Center & Research Institute, Tampa, FL, USA. ^53^ Institute of Cancer and Genomic Sciences, University of Birmingham, Birmingham, UK. ^54^ Department of Women's and Children's Health, Karolinska Institutet, Stockholm, Sweden. ^55^ Department of Biochemistry and Molecular Medicine, University of California Davis, Davis, CA, USA. ^56^ School of Medicine and Public Health, University of Newcastle, Callaghan, New South Wales, Australia. ^57^ Harvard T.H. Chan School of Public Health, Boston, MA, USA. ^58^ Dana-Farber Cancer Institute, Boston, MA, USA. ^59^ Department of Gynaecology and Obstetrics, University Hospital Erlangen, Friedrich-Alexander University Erlangen-Nuremberg, Comprehensive Cancer Center Erlangen-EMN, Erlangen, Germany. ^60^ Center for Cancer Prevention (CPO-Peimonte), Turin, Italy. ^61^ Human Genetics Foundation (HuGeF), Turino, Italy. ^62^ Department of Obstetrics and Gynecology, School of Medicine and Public Health, University of Wisconsin, Madison, WI, USA. ^63^ Department of Population and Quantitative Health Sciences, Case Western Reserve University, Cleveland, OH, USA. ^64^ Division of Molecular Medicine, Pathology North, John Hunter Hospital, Newcastle, New South Wales, Australia. ^65^ Clinical Genetics, Karolinska Institutet, Stockholm, Sweden. ^66^ Population Health Department, QIMR Berghofer Medical Research Institute, Brisbane, Queensland, Australia. ^67^ Department of Health Sciences Research, Division of Biomedical Statistics and Informatics, Mayo Clinic, Rochester, MN, USA. ^68^ State Key Laboratory of Oncogene and Related Genes & Department of Epidemiology, Shanghai Cancer Institute, Renji Hospital, Shanghai Jiaotong University School of Medicine, Shanghai, China.

**Endometrial Cancer Association Consortium Acknowledgements**

The authors thank the many individuals who participated in this study and the numerous institutions and their staff who supported recruitment.

The iCOGS and OncoArray endometrial cancer analysis were supported by NHMRC project grants [ID#1031333 & ID#1109286] Funding for the iCOGS infrastructure came from: the European Community's Seventh Framework Programme under grant agreement no 223175 [HEALTH-F2-2009-223175] [COGS], Cancer Research UK [C1287/A10118, C1287/A 10710, C12292/A11174, C1281/A12014, C5047/A8384, C5047/A15007, C5047/A10692, C8197/A16565], the National Institutes of Health [CA128978] and Post-Cancer GWAS initiative [1U19 CA148537, 1U19 CA148065 and 1U19 CA148112 - the GAME-ON initiative], the Department of Defence [W81XWH-10-1-0341], the Canadian Institutes of Health Research [CIHR] for the CIHR Team in Familial Risks of Breast Cancer, Komen Foundation for the Cure, the Breast Cancer Research Foundation, and the Ovarian Cancer Research Fund. OncoArray genotyping of ECAC cases was performed with the generous assistance of the Ovarian Cancer Association Consortium (OCAC). We particularly thank the efforts of Cathy Phelan. The OCAC OncoArray genotyping project was funded through grants from the US National Institutes of Health (CA1X01HG007491-01, U19-CA148112, R01-CA149429 and R01-CA058598); Canadian Institutes of Health Research (MOP-86727); and the Ovarian Cancer Research Fund. CIDR genotyping for the Oncoarray was conducted under contract 268201200008I. OncoArray genotyping of the BCAC controls was funded by Genome Canada Grant GPH-129344, NIH Grant U19 CA148065, and Cancer UK Grant C1287/A16563.

Stage 1 and stage 2 case genotyping was supported by the NHMRC [ID#552402, ID#1031333]. Control data were generated by the Wellcome Trust Case Control Consortium (WTCCC), and a full list of the investigators who contributed to the generation of the data is available from the WTCCC website. We acknowledge use of DNA from the British 1958 Birth Cohort collection, funded by the Medical Research Council grant G0000934 and the Wellcome Trust grant 068545/Z/02 - funding for this project was provided by the Wellcome Trust under award 085475. NSECG was supported by the EU FP7 CHIBCHA grant, Wellcome Trust Centre for Human Genetics Core Grant 090532/Z/09Z, and CORGI was funded by Cancer Research UK. We thank Nick Martin, Dale Nyholt and Anjali Henders for access to GWAS data from QIMR Controls. Recruitment of the QIMR controls was supported by the NHMRC. The University of Newcastle, the Gladys M Brawn Senior Research Fellowship scheme, The Vincent Fairfax Family Foundation, the Hunter Medical Research Institute and the Hunter Area Pathology Service all contributed towards the costs of establishing the Hunter Community Study. The WHI program is funded by the National Heart, Lung, and Blood Institute, the US National Institutes of Health and the US Department of Health and Human Services (HHSN268201100046C, HHSN268201100001C, HHSN268201100002C, HHSN268201100003C, HHSN268201100004C and HHSN271201100004C). This work was also funded by NCI U19 CA148065-01. This research has been conducted using the UK Biobank Resource under applications 5122 and 9797.

ANECS recruitment was supported by project grants from the NHMRC [ID#339435], The Cancer Council Queensland [ID#4196615] and Cancer Council Tasmania [ID#403031 and ID#457636]. SEARCH recruitment was funded by a programme grant from Cancer Research UK [C490/A10124]. The Bavarian Endometrial Cancer Study (BECS) was partly funded by the ELAN fund of the University of Erlangen. The Hannover-Jena Endometrial Cancer Study was partly supported by the Rudolf Bartling Foundation. The Leuven Endometrium Study (LES) was supported by the Verelst Foundation for endometrial cancer. The Mayo Endometrial Cancer Study (MECS) and Mayo controls (MAY) were supported by grants from the National Cancer Institute of United States Public Health Service [R01 CA122443, P30 CA15083, P50 CA136393, and GAME-ON the NCI Cancer Post-GWAS Initiative U19 CA148112], the Fred C and Katherine B Andersen Foundation, the Mayo Foundation, and the Ovarian Cancer Research Fund with support of the Smith family, in memory of Kathryn Sladek Smith. MoMaTEC received financial support from a Helse Vest Grant, the University of Bergen, Melzer Foundation, The Norwegian Cancer Society (Harald Andersens legat), The Research Council of Norway and Haukeland University Hospital. The Newcastle Endometrial Cancer Study (NECS) acknowledges contributions from the University of Newcastle, The NBN Children’s Cancer Research Group, Ms Jennie Thomas and the Hunter Medical Research Institute. RENDOCAS was supported through the regional agreement on medical training and clinical research (ALF) between Stockholm County Council and Karolinska Institutet [numbers: 20110222, 20110483, 20110141 and DF 07015], The Swedish Labor Market Insurance [number 100069] and The Swedish Cancer Society [number 11 0439]. The Cancer Hormone Replacement Epidemiology in Sweden Study (CAHRES, formerly called The Singapore and Swedish Breast/Endometrial Cancer Study; SASBAC) was supported by funding from the Agency for Science, Technology and Research of Singapore (A*STAR), the US National Institutes of Health and the Susan G. Komen Breast Cancer Foundation.

The Nurses’ Health Study (NHS) is supported by the NCI, NIH Grants Number UM1 CA186107, P01 CA087969, R01 CA49449, 1R01 CA134958, and 2R01 CA082838. The authors thank the participants and staff of the Nurses’ Health Study for their valuable contributions as well as the following state cancer registries for their help: AL, AZ, AR, CA, CO, CT, DE, FL, GA, ID, IL, IN, IA, KY, LA, ME, MD, MA, MI, NE, NH, NJ, NY, NC, ND, OH, OK, OR, PA, RI, SC, TN, TX, VA, WA, WY. The authors assume full responsibility for analyses and interpretation of these data. The authors also thank Channing Division of Network Medicine, Department of Medicine, Brigham and Women’s Hospital and Harvard Medical School. Finally, the authors also acknowledge Pati Soule and Hardeep Ranu for their laboratory assistance. The Connecticut Endometrial Cancer Study was supported by NCI, NIH Grant Number RO1CA98346. The Fred Hutchinson Cancer Research Center (FHCRC) is supported by NCI, NIH Grant Number NIH RO1 CA105212, RO1 CA 87538, RO1 CA75977, RO3 CA80636, NO1 HD23166, R35 CA39779, KO5 CA92002 and funds from the Fred Hutchinson Cancer Research Center. The Multiethnic Cohort Study (MEC) is supported by the NCI, NHI Grants Number CA54281, CA128008 and 2R01 CA082838. The California Teachers Study (CTS) is supported by NCI, NIH Grant Number 2R01 CA082838, R01 CA91019 and R01 CA77398, and contract 97-10500 from the California Breast Cancer Research Fund. The Polish Endometrial Cancer Study (PECS) is supported by the Intramural Research Program of the NCI. The Prostate, Lung, Colorectal, and Ovarian Cancer Screening Trial (PLCO) is supported by the Extramural and the Intramural Research Programs of the NCI.
